# Home and Epigenome: DNA Methylation as a Link Between Poor Housing Quality and Depressive Symptoms

**DOI:** 10.1101/2024.10.21.24315867

**Authors:** Faye Sanders, Vilte Baltramonaityte, Alexandre A. Lussier, Andrew D.A.C. Smith, Erin C Dunn, Esther Walton

**Affiliations:** Department of Psychology, University of Bath, Bath, UK; Psychiatric and Neurodevelopmental Genetics Unit, Center for Genomic Medicine, Massachusetts General Hospital, Boston, MA; Department of Psychiatry, Harvard Medical School, Boston, MA; Stanley Center for Psychiatric Research, The Broad Institute of Harvard and MIT, Cambridge, MA; Mathematics and Statistics Research Group, University of the West of England, Bristol, UK; Department of Sociology, College of Liberal Arts, Purdue University, West Lafayette, IN, United States

**Author notes:** Corresponding author: Esther Walton, Department of Psychology, University of Bath, Claverton Down, Bath, BA2 7AY,.

**Keywords:** depression, ALSPAC, housing quality, DNA methylation

## Abstract

Poor housing quality associates with risk for depression. However, previous research often lacks consideration of socioeconomic status (SES) baseline depressive symptoms and biological processes, leading to concerns of confounding and reverse causation.

In a sample of up to 9,566 adults, we investigated cross-sectional and longitudinal associations between housing quality (assessed at age 28, 1- and 2-year follow-ups) and depressive symptoms (at four intervals between enrolment and 18-year follow-up). In sub-samples (n=871, n=731), we investigated indirect effects via DNA methylation.

Poor housing quality associated with depressive symptoms cross-sectionally (beta range: 0.02 to 0.06, all *p*-values<.023) after controlling for SES and other factors. Longitudinally, this association persisted at the ∼2-year, but not the ∼18-year follow-up period. Indirect effects linked to genes related to aging, obesity, and brain health.

These results highlight poor housing quality as a risk factor for depression and the potential role of DNA methylation in this association.

## Introduction

With individuals spending large amounts of time at home ^1^, consistent and potentially long-lasting associations between poor housing quality and poor mental health have been reported ^2,3^. For example, a systematic review on poor housing and mental health found positive associations in seven out of eight studies, whereby people experiencing poor housing conditions were nearly eight times more likely to have depression ^3^. The large effect sizes highlight the importance of investigating housing as a means to improve population mental health. Given that the association between poor housing quality and depression become greater as we age ^4,5^, the need to study this relationship across adulthood is critical.

However, current research on the relationship between the physically-built environment (e.g., housing quality) faces five challenges. First, whilst great attention has been paid to disentangling associations between macro features of urbanicity, including population density and the built environment, and mental health outcomes ^6–8^, less focus has been put on smaller-scale features, including housing quality, which may be in some aspects easier to intervene on. Second, research detecting associations between housing quality and mental health has largely focused on adults living in deprived neighbourhoods ^3,9^. As participants are often sampled from lower-income housing developments ^10,11^, it remains unclear whether these associations are specific to housing quality, or due to confounding effects of lower socioeconomic status (SES) ^12^.

Third, even when studies considered a larger socioeconomic spectrum, SES was often operationalized through single proxy measures including income ^13^. A more fine-grained assessment of SES in housing quality and health studies is critical to ensure that broader aspects of SES, including education, family contexts, occupation, and life experiences ^14,15^ are captured, in addition to income inequalities.

Fourth, baseline mental health is often omitted in previous research, opening up the possibility for reverse causation. For example, the ‘social drift hypothesis’ of urbanicity ^8,16^ posits that individuals with a greater risk of experiencing mental health difficulties (due to genetic or environmental factors) are more likely to move to or stay in environments associated with poor mental health outcomes. So far, studies testing this reverse association have focused on macro levels of urbanicity, including population density ^17^. Thus, our understanding of how this social drift hypothesis applies to smaller-scale measures, such as housing quality, remain limited.

Finally, we do not yet have a good understanding of the biological processes through which housing quality might associate with mental health outcomes. Furthering this understanding is particularly important as, similarly to neighbourhood characteristics, it may be that these associations are not limited to depression and have important implications for the understanding of mental health more broadly ^18^.

Complex traits, including depression, arise from the interplay of both genetic and environmental factors ^19,20^ for which epigenetic processes may be a proxy. One of the most studied epigenetic mechanisms is DNA methylation (DNAm), which represents the addition or removal of methyl groups to cytosine-guanine (CpG) base pairs on the genome ^21^. DNAm has been associated with many environmental exposures such as neighbourhood green space ^22^ and neighbourhood disadvantage ^23,24^. Additionally, a smaller body of evidence has found that DNAm might mediate associations between these large-scale environmental exposures and poor mental health outcomes ^25^. Yet, it remains unclear whether associated factors drive these associations. For example, the association between neighbourhood green space and DNAm ^22^ could be driven by confounding (e.g., people living near green spaces are also more likely to exercise). Hence, studying a component of the physically-built environment that is less directly linked to exercise opportunities (e.g., housing quality, rather than neighbourhood green spaces; ^26^, may provide further clarity.

To address these challenges, we investigated if 1) poor housing quality predicts depressive symptoms in adulthood whilst controlling for a comprehensive battery of SES measures and history of depression, in a broad population-based sample; and 2) DNAm in early and mid-adulthood partly explains these associations.

## Results

### Descriptive Statistics

All participants were female and aged between 15-44 years at enrolment (**Table 1**). Twenty-two percent had a university degree and an additional 30% completed their A-levels. The average number of times moved in the last 5 years at the beginning of the study was 1.65 (*SD* = 1.75).

**Table 1.** Study descriptives for 9,566 women who had completed questionnaire items for their depressive symptoms at enrolment and/or ∼18-year follow-up (imputed data). ^1^ *Depressive symptoms measured at enrolment, and at 1- and 2-year follow-up used the Crown Crisp Experiential Index. Depressive symptoms measured at ∼18-year follow-up used the 36-Item Short Form Survey mental health scale*. *^2^ Ethnicity (e.g., black/Caribbean, black/African, black/other, Indian, Pakistani, Bangladeshi, Chinese, any other ethnic group) was self-reported at study enrolment. SD = standard deviation; SES = socioeconomic status*.

|  | Enrolment (Mean<br>age 28) | 1-yr follow-up | 2-yr follow-up | 18-yr follow-up |
| --- | --- | --- | --- | --- |
| Poor Housing Quality |  |  |  |  |
| Mean (SD) | 3.44 (0.34) | 1.90 (0.22) | 2.97 (0.37) | NA |
| Min, Max | 1.21, 4.81 | 1.01, 3.47 | 1.05, 4.27 |  |
| SES Risk Factors |  |  |  |  |
| Mean (SD) | 1.63 (0.88) | 2.02 (0.93) | 1.96 (0.94) | NA |
| Min, Max | 0.60, 5.60 | 0.80, 6.20 | 0.80, 6.80 |  |
| Familial Depressive History |  |  |  |  |
| Both parents | 5.27% |  | NA |  |
| One parent | 20.64% |  |  |  |
| None | 74.10% |  |  |  |
| Housing Instability |  |  |  |  |
| Mean (SD) | 1.65 (1.75) | 1.40 (1.71) | 0.23 (0.55) | NA |
| Min, Max | 0.00, 28.00 | 0.00, 22.00 | 0.00, 8.00 |  |
| Poor Neighbourhood Quality |  |  |  |  |
| Mean (SD) | 4.74 (2.19) | NA | 3.89 (3.52) | NA |
| Min, Max | 1.00, 13.00 |  | 0.00, 22.00 |  |
| Depressive Symptoms <sup>1</sup> |  |  |  |  |
| Mean (SD) | 4.30 (3.02) | 3.30 (2.97) | 2.91 (2.68) | 14.70 (4.38) |
| Min, Max | 0.00, 16.00 | 0.00, 16.00 | 0.00, 14.00 | 3.00, 25.00 |
| **Age (years)** |  |  |  |  |
| Mean (SD) | 28.36 (4.67) | 29.46 (4.68) | 29.99 (4.73) | 47.71 (4.72) |
| Min, Max | 15.00, 44.00 | 16.00, 45.00 | 15.00, 63.00 | 34.00, 63.00 |
| **Ethnicity<sup>2</sup>** |  |  |  |  |
| White | 98.12% |  |  |  |
| Non-White | 1.88% |  | NA |  |
| **Smoking at enrolment** |  |  |  |  |
| Never | 51.94% |  |  |  |
| Until pregnancy | 29.70% |  | NA |  |
| During pregnancy | 18.35% |  |  |  |
| **Smoking at 18-yr follow-up** |  |  |  |  |
| 0 per day |  |  |  | 53.38% |
| 1-15 per day |  | NA |  | 29.99% |
| 20+ per day |  |  |  | 16.63% |

The DNAm subsamples were much smaller (due to DNAm data only being collected in smaller sample), with some small differences between each subsample to the overall cohort for some measures (see SM Table S1). For example, contextual SES risk and smoking rates were slightly higher in the full sample than in the DNAm subsamples.

#### Step 1: Cross-sectional associations between poor housing quality and depressive symptoms

Cross-sectionally, poor housing quality was significantly associated with increased depressive symptoms at all three timepoints (study enrolment, 1- and 2-year follow-ups; β range: 0.02 to 0.06, all p-values<.023). In all three models, this association held whilst controlling SES risk factors, familial depressive history, poor neighbourhood quality, and housing instability, and additionally baseline depressive symptoms at enrolment in model 2 and 3 (**Table 2**).

**Table 2.** Cross-sectional and longitudinal standardised regression results for association between poor housing quality and depressive symptoms.

| Model | Exposure | Standardised Estimate |  | Adjusted R<sup>2</sup> | sr<sup>2</sup> | p value |
| --- | --- | --- | --- | --- | --- | --- |
|  |  | Adjusted<br>[95% CI] | Unadjusted<br>[95% CI] |  |  |  |
| Cross-sectional at study enrolment | Poor Housing Quality (at study enrolment) | 0.024<br>[0.03, 0.45] | 0.112<br>[0.09,0.13] | .101 | .005 | .018 |
| Cross-sectional at 1-year follow-up | Poor Housing Quality (at 1- year follow-up) | 0.059<br>[0.04, 0.08] | 0.092<br>[0.07, 0.11] | .297 | .006 | < .0001 |
| Cross-sectional at 2-year follow-up | Poor Housing Quality (at 2- year follow-up) | 0.020<br>[0.002, 0.04] | 0.077<br>[0.06, 0.10] | .271 | .002 | .023 |
| Longitudinal ~2y follow-up | Poor Housing Quality (at study enrolment) | 0.032<br>[0.02, 0.05] | 0.110<br>[0.09, 0.13] | .244 | .005 | < .001 |
| Longitudinal<br>~18y follow-up | Poor<br>Housing<br>Quality (at<br>1- and 2-<br>year follow-ups) | -0.006<br>[-0.02, 0.02] | 0.024<br>[0.004, 0.04] | .037 | .0002 | .956 |

Depressive symptoms were measured using the Crown Crisp Experiential Index (at study enrolment, 1- and 2- year follow-ups) and the 36-Item Short Form Survey (at ~18y follow-up). Standardised estimates are measured on a scale of increase in standard deviations of depressive symptoms per standard deviation increase in exposure. Therefore, standardised estimates can be interpreted on the same scale as a correlation coefficient. Adjusted estimates refer to models including covariates, while unadjusted estimates are covariates-uncorrected. In cross-sectional models (study enrolment, 1- and 2-year follow-ups) and in the 18-year longitudinal follow-up, covariates included socioeconomic status risk factors, familial depressive history, housing instability, age at depressive symptoms. Baseline depressive symptoms were additionally included as a covariate in 1- and 2- year follow-up cross-sectional models and the 18-year longitudinal follow-up. In cross-sectional models (study enrolment and 2-year follow-up) and in longitudinal 2- and 18-year follow-ups, poor neighbourhood quality was an additional covariate. Adjusted R2 value refers to the R2 estimate which is penalised based on the number of predictors entered into the model. sr2 = semi-partial correlation coefficient.

#### Step 2 – Longitudinal analyses of associations between poor housing quality and follow-up depressive symptoms

Poor housing quality experienced during early adulthood (at study enrolment) significantly associated with depressive symptoms at 2-year follow-up (β=.03, SE=.01, sr^2^=.005, *p*<.001; **Table 2 and Figure 1A**). Poor housing quality experienced during early adulthood (at 1- and 2-year follow-ups) did not significantly associate with depressive symptoms measured 18 years later (β=-0.01, SE=.01, sr^2^=.0002, *p*=.956, **Table 2 and Figure 1B**).

**Figure 1.**
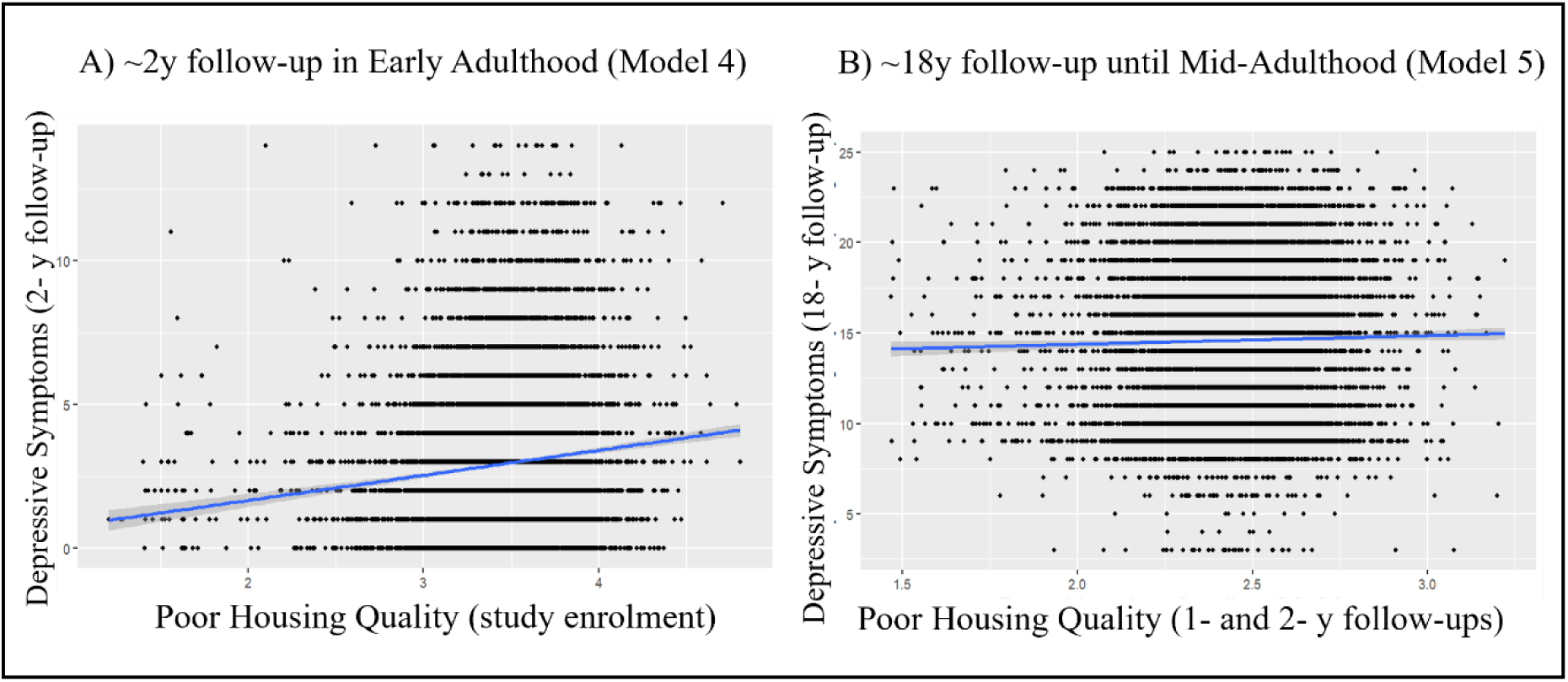
Longitudinal associations between A) poor housing quality at study enrolment and depressive symptoms at 2-year follow-up, and B) between poor housing quality at 1- and 2-year follow-ups and depressive symptoms at 18-year follow-up.

#### Step 3 – DNAm Indirect Effects Analyses

In model 6, out of the 425 CpG sites identified as *potential* intermediary variables from sure independence screening (SM section 1.3), eight showed evidence of indirect effects in the association between poor housing quality (at study enrolment) and depressive symptoms two years later (**Table 3**). Poor housing quality was associated with decreased methylation of one CpG and increased methylation of seven CpGs (SM Figures 1), which, in turn, were associated with decreased and increased depressive symptoms respectively.

**Table 3.**
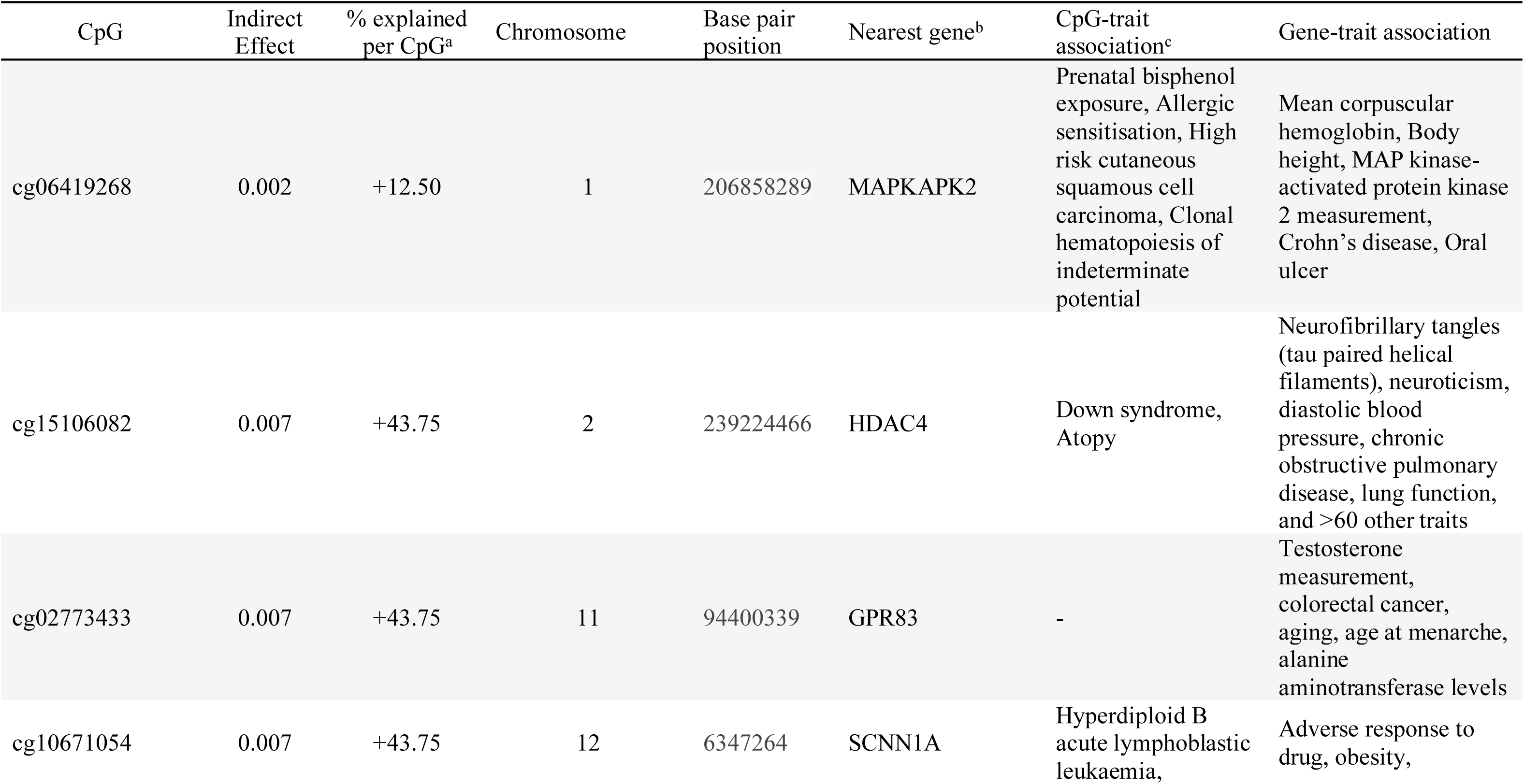
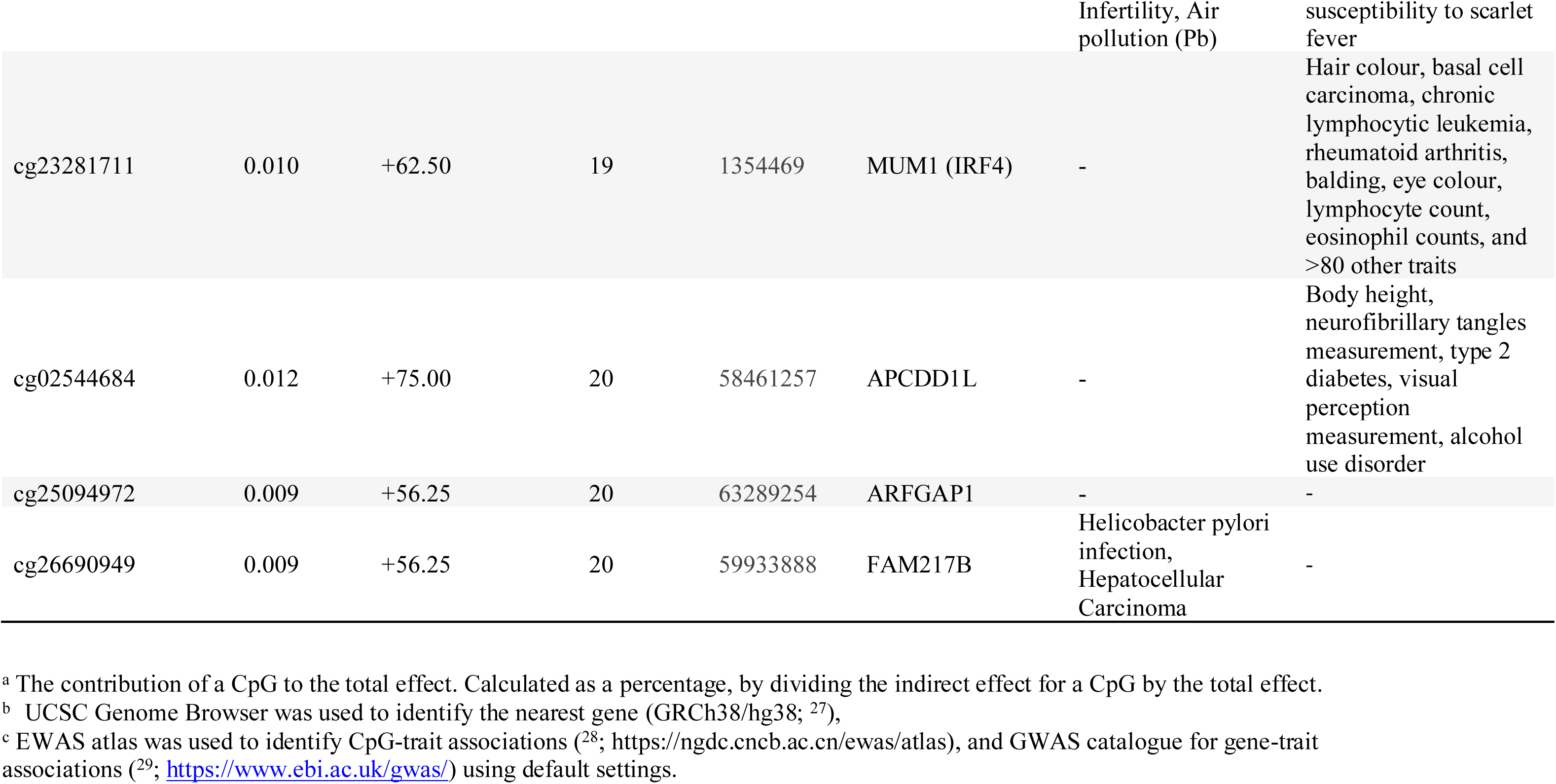
CpGs explaining the association between poor housing quality in early adulthood and depressive symptoms at ∼2y follow-up (n = 871).

In model 7, 364 CpGs were identified as potential intermediary variables in the association between poor housing quality (at 1- and 2-year follow-up) and depressive symptoms (at 18-year follow-up). However, in line with a non-significant total effect, none of these CpGs showed evidence of indirect effects.

Further, the indirect effects, across all CpGs, was positive (β=0.002 to 0.012). Poorer housing quality was associated with increased depressive symptoms via DNAm (**Table 4**, SM Figure 2).

**Figure 2.**
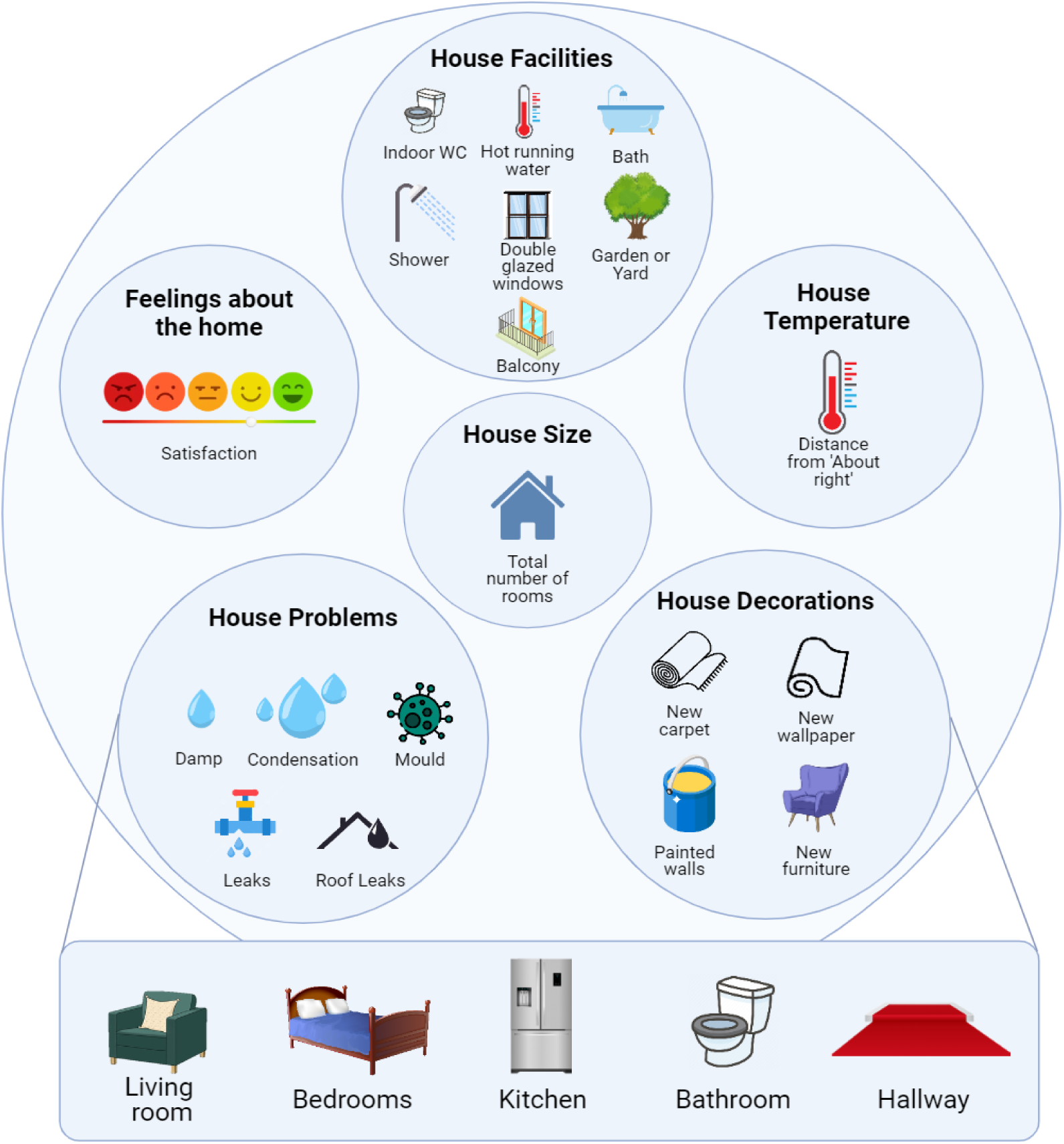
List of variables included in the ‘Poor Housing Quality’ score.

**Table 4.** DNAm indirect effects summary for the association between poor housing quality in early adulthood and depressive symptoms at the ∼2y follow-up (n = 871, model 6).

| N | Standardized Total Effect <sup>a</sup> |  |  | Total Indirect Effect | Direct Effect <sup>b</sup> |
| --- | --- | --- | --- | --- | --- |
| | $\beta$ | SE | p | $\beta$ | $\beta$ |
| 871 | 0.016 | 0.031 | 0.566 | 0.063 | -0.047 |

<sup>a</sup> The total effect of poor housing quality on depressive symptoms in DNAm subsample ( $n = 871$ ), standardized so the estimate can be interpreted as the correlation between poor housing quality at study enrolment and depressive symptoms at 2-year follow-up.
<sup>b</sup> Direct effect is the effect of poor housing quality on depressive symptoms independent of any indirect effects.

## Discussion

We investigated the relationship between poor housing quality and depressive symptoms and the indirect effect of DNAm in potentially explaining this relationship. Our results showed that 1) poor housing quality predicted depressive symptoms even after controlling for key sociodemographic confounders, both cross-sectionally and longitudinally but only at the 2-year and not the 18-year follow-up; and 2) DNA methylation of selected CpG sites partly explained this association during a ∼2y follow-up, but not ∼18y follow-up period. We highlight and discuss five key points.

### Cross-sectional and longitudinal associations between poor housing quality and depressive symptoms

First, we found that poor housing quality associated with higher depressive symptoms both cross-sectionally and longitudinally at the 2-year follow-up, but not the 18-year follow-up. This finding is consistent with previous research ^2^, which has reported weakening associations between poor housing and mental health over a three-year period ^2^. The lack of association at the 18-year follow-up may be because participants no longer lived at the same address, and/or their housing conditions had since improved. Equally, even if participants’ housing conditions remained the same, other factors may have become stronger predictors of mental health at this timepoint. Our findings suggest that poor housing conditions may have acute negative effects on mental health. However, further research that includes assessments of housing conditions at later time points is needed to better understand these relationships between housing quality and mental health later in life.

### Importance of micro features of urbanicity

Second, consistent cross-sectional associations between poor housing quality and depressive symptoms highlight the importance of considering micro features of urbanicity, including housing quality. A large focus of previous research has focussed on the role of macro features, such as population density, in predicting health outcomes ^7,8^. Although we were able to replicate the importance of macro features (e.g. neighbourhood quality), housing quality was independently predictive of depressive symptoms over and above these macro feature effects. Our findings suggest that considering both individual and population-level features of urbanicity may better predict poor mental health outcomes than when used in isolation.

### Role of SES

Third, our findings suggest the associations between poor housing quality and depressive symptoms are not fully confounded by or restricted to low SES contexts. As previous research was predominantly based on deprived samples ^10,11^, it was still unclear whether these associations are also present in population-based samples across a wider SES spectrum. Whilst housing quality and SES are interconnected, our findings show poor housing quality might be a risk factor for depression over a broader range – and partially independent of – SES. This finding is further supported by the depth of our SES risk factors score, which covered education, income, and homelessness, in comparison to previous literature, which used single proxy measures of SES such as occupational status ^13^.

### Poor housing quality as a risk factor for depressive symptoms

Fourth, associations between poor housing quality and depressive symptoms were present even when controlling for baseline depressive symptoms. These results suggest that poor housing quality might be an antecedent for depressive symptoms, rather than vice versa. Although we cannot draw any causal conclusions, improving housing conditions could potentially help reduce depressive symptoms, further emphasizing the role of socio-environmental factors in mental health interventions.

### The role of DNAm in associations between poor housing quality and depression

Fifth, we found some evidence that DNAm levels at eight CpGs might partly explain the associations between poor housing quality and depressive symptoms in early adulthood. This finding provides further support for potential biological correlates of the associations between risk exposures and health outcomes ^30^. Most of these eight CpGs have been previously associated with various health phenotypes. For example, cg15106082 (nearest gene: *HDAC4*) has been linked to neuroticism ^31^, neurofibrillary tangles ^32^ and lung function ^33^. Cg02773433 (nearest gene: *GPR83* gene) has been linked to aging ^34^, cg10671054 (nearest gene: *SCNN1A*) to obesity ^35^ and air pollution ^36^, and cg02544684 (nearest gene: *APCDD1L*) to neurofibrillary tangles ^32^. Out of these eight CpGs, cg02544684 explained the largest proportion of variance in the association between housing quality and depressive symptoms (∼75% of the total effect). All these phenotypes (neuroticism, neurofibrillary tangles, lung function, aging, obesity) and air pollution have been associated with depression in previous studies, reflecting the biological plausibility of the identified CpGs ^31–33,35,36^. One of the eight CpGs (cg25094972; explaining approximately 52% of the total effect) and genes (*ARFGAP1*) had no known trait associations, suggesting a potentially novel depression pathway.

### Limitations

Our findings must be considered alongside its limitations. Indirect associations do not reflect causality. Despite our efforts to minimise reverse causation, future research may wish to triangulate our findings using (quasi-)experimental designs. Additionally, DNAm was measured in blood samples, limiting our ability to draw conclusions regarding brain-based processes. Lastly, indirect effects were generally small and short-term, and tested in a much smaller sample than in the main analyses. This change in sample size may have led to some unexpected findings. For example, in the epigenetic subsample, we observed opposite signs for the total indirect effect and the direct effect between poor housing quality and depressive symptoms. This unexpected direction of effect (e.g. better housing quality - poorer mental health) have been reported in previous research identifying that in some cases improvements to housing conditions can worsen mental health due to disturbances to social networks ^37^. However, in the overall sample (possibly due to better power) we observed a significant and adverse total effect, indicating that efforts should be made to reduce poor housing quality to improve depressive symptoms.

## Conclusions

To conclude, we found poor housing quality was associated with increased depressive symptoms, and showed that DNAm might partially explain this relationship in early adulthood. Our findings highlight the importance of housing quality for mental well-being, and the urgency to improve housing conditions for both current and future population health.

## Methods

### Participants

Participants were mothers from the Avon Longitudinal Study of Parents and Children (ALSPAC) cohort ^38–40^. Pregnant women resident in Avon, UK with expected dates of delivery 1st April 1991 to 31st December 1992 were invited to take part in the study and the initial number of pregnancies enrolled was 14,541. Of the original 14,541 initial pregnancies, 14,203 unique mothers were initially enrolled in the study. Additional phases of recruitment provided a total of 14,833 unique women. We selected mothers who had completed questionnaire items for their housing circumstances at study enrolment or, 1- or 2-year follow-ups, leaving up to 9,566 mothers available for analysis. Missing data for the exposure, covariates, and outcomes were imputed (see supplementary materials section 1.1 for further information).

DNAm measures were available in mothers who participated in the Accessible Resource for Integrated Epigenomic Studies (ARIES) ^41^, comprising a subset of n=871 participants for analysis at study enrolment and a subset of n=731 participants 18 years later.

Ethical approval for the study was obtained from the ALSPAC Ethics and Law Committee and the Local Research Ethics Committees and Department of Psychology at the University of Bath. Informed consent for the use of data collected via questionnaires and clinics was obtained from participants following the recommendations of the ALSPAC Ethics and Law Committee at the time. Consent for biological samples has been collected in accordance with the Human Tissue Act (2004). The study website (https://www.bristol.ac.uk/alspac/researchers/our-data/) contains details of all data available through a fully searchable data dictionary and variable search tool.

## Measures

### Poor Housing Quality

General housing data (e.g., size, temperature, facilities, decorations, problems, and feelings towards the home; Figures 2) was collected using self-report questionnaires completed by the mothers at study enrolment (age 28), and at 1- and 2-year follow-ups. House size was measured via the total number of rooms in the property. Facilities were measured by asking if the property had access to the following features or not, ‘Hot running water’, ‘Indoor WC’, ‘Bath’, ‘Shower’, ‘Garden or Yard’, ‘Balcony’, and ‘Double glazed windows’. Decorations measured whether the living room, bedrooms, kitchen, and other rooms had modifications made to them in the last year including new paint, wallpaper, carpet, or furniture. Temperature was measured via ‘During the coldest time of year, describe the temperature in your bedrooms and living room’ with responses including ‘Very cold’, ‘Cold’, ‘0/About right’, ‘Warm’, and ‘Very warm’. Temperature was coded as deviation from ‘0/About right’, with greater deviations contributing to poorer housing quality. Problems measured both the presence and severity of leaks, damp, condensation, and mould in the home with the following responses, ‘Very serious’, ‘Fairly serious’, ‘Not serious’, ‘No leak/damp/condensation’. Lastly, feelings towards the home were measured by asking ‘Taking everything into account, which of the following best describes your feelings about your home?’ with responses including ‘Very dissatisfied’, ‘Dissatisfied’, ‘Fairly satisfied’, and ‘Very satisfied’. Questionnaires contained items from validated and reliable indicators of housing quality ^10^ and from the UK ‘Decent Housing Standard’ ^42^. House size, facilities, decorations were reverse coded, to ensure higher scores indicated poorer housing quality, and then averaged together with the remaining general housing data to produce a ‘poor housing quality’ score.

### Depressive Symptoms

Depressive symptoms were measured at study enrolment, 1- and 2-year follow-ups using the Crown Crisp Experiential Index (CCEI) ^43^. The CCEI is a commonly-implemented measure of depression which comprehensively assesses features of depression including anhedonia and sadness. The CCEI consisted of 10 items such as, ‘I have felt sad or miserable’ and ‘I have been so unhappy that I have been crying’, with responses including ‘Yes, most of the time’, ‘Yes, quite often’, ‘Not very often/Only occasionally’ and ‘No, not at all/No, never’. Scores can range from 0 to 30. At 18 years follow-up, depressive symptoms were measured via the 36-Item Short Form Survey (SF-36) Mental Health subscale, a valid and reliable measure of mental health ^44^ and a strong predictor of depressive symptoms ^45^. Both the CCEI and SF-36 can distinguish well between groups of depressed and non-depressed patients ^46,47^. The subscale included 9 questions such as, ‘How much during the last 2 weeks have you felt so down in the dumps nothing would cheer you up’ with responses as ‘All of the time’, ‘Most of the time’, ‘A good bit of the time’, ‘Some of the time’, ‘A little of the time’ and ‘None of the time’ (score range: 0-45).

### Housing Instability

Housing instability was measured by asking mothers for the number of times moving house in the last 5 years, measured at study enrolment and 1-year follow-up, and the number of times of moving home since the child was 8 months, at 2-year follow-up. Housing instability was coded so that higher scores indicated moving home more frequently (i.e., greater housing instability).

### Poor Neighbourhood Quality

Neighbourhood quality was self-reported at study enrolment measuring liveliness, friendliness, noise, cleanliness, attractiveness, and pollution/dirt with responses including ‘Yes, usually’, ‘Yes, sometimes’, and ‘No not at all’. Neighbourhood quality was also self-reported at 2-year follow-up measuring pollution/dirt and crime with responses including ‘Serious problem’, ‘Minor problem’, ‘Not a problem’, and ‘No opinion’. Higher scores indicated worse neighbourhood quality. Further details can be seen in the supplementary materials (SM) section 1.2.

### Familial Depressive History

Familial depressive history (e.g., known lifetime experience of depression or consistent low mood in the mothers’ parents) was reported by the mothers at study enrolment.

### SES Risk Factors

SES risk factors scores – comprised of mother’s education, experience of reduced income, losing a job, becoming homeless, and financial difficulties – were measured at study enrolment, 1- and 2-year follow-ups. Education was coded, in order of highest to lowest SES risk factors, as ‘CSE’, ‘Vocational’, ‘O level’, ‘A level’, and ‘Degree’. Experiences of reduced income, becoming homeless and financial difficulties, in order of highest to lowest SES risk factors, were coded as ‘Affected a lot’, ‘Moderately affected’, ‘Mildly affected’, ‘No effect at all’, and ‘Did not happen’.

### DNA methylation (DNAm)

Mothers’ peripheral blood samples were collected at study enrolment and 18 years later. Sodium bisulfite conversion was used on 500 ng of DNA using the EZ-DNAm kit (Zymo Research, Orange, CA, USA) and scanned using the Illumina iScan (software version 3.3.28). DNAm levels of CpG sites was measured using the Illumina HumanMethylation450 BeadChip (Illumina, USA), which covers approximately 485,000 CpG sites across the epigenome.

Normalization of DNAm data was performed using the ‘*Meffil’* R package ^48^. Non-specific probes were removed prior to analysis to improve the quality of DNAm data ^49^, leaving 420,752 probes for analysis. Prior to running indirect effects analyses (see next section), DNAm data were residualised for cell type, smoking, batch, and age at DNAm collection.

### Cell Type Proportions

Estimated counts of cell type (B cell, CD4T, CD8T, granulocyte, monocyte, natural killer cell, nucleated red blood cell) were derived using the blood gse35069 complete panel ^50^ available in the *Meffil* package ^48^, for DNAm measured at both study enrolment and at 18-year follow up.

### Smoking Exposure

Smoking behaviour was self-reported at enrolment in the study (‘smoked regularly in her last two months of pregnancy’), and 18 years later (number of cigarettes smoked per day on average).

### Batch Effects

Batch at both DNAm measurements (study enrolment and 18 years later) were controlled for by performing surrogate variable analysis in *meffil* and extracting 10 surrogate variables that were not associated with the relevant outcomes of depressive symptoms (study enrolment and 18 years later respectively).

## Statistical Analyses

This current study was divided into three steps (Figure 3 **and SM** Figure 1). First, we performed three cross-sectional analyses (at study enrolment and at 1- and 2-year follow-ups in regression models 1, 2 and 3 respectively) testing the association between poor housing quality and depressive symptoms at each timepoint, whilst accounting for SES risk factors, poor neighbourhood quality (in models 1 and 3), housing instability, and familial depressive history. In models 2 and 3 we also accounted for baseline depressive symptoms (at study enrolment).

**Figure 3.**
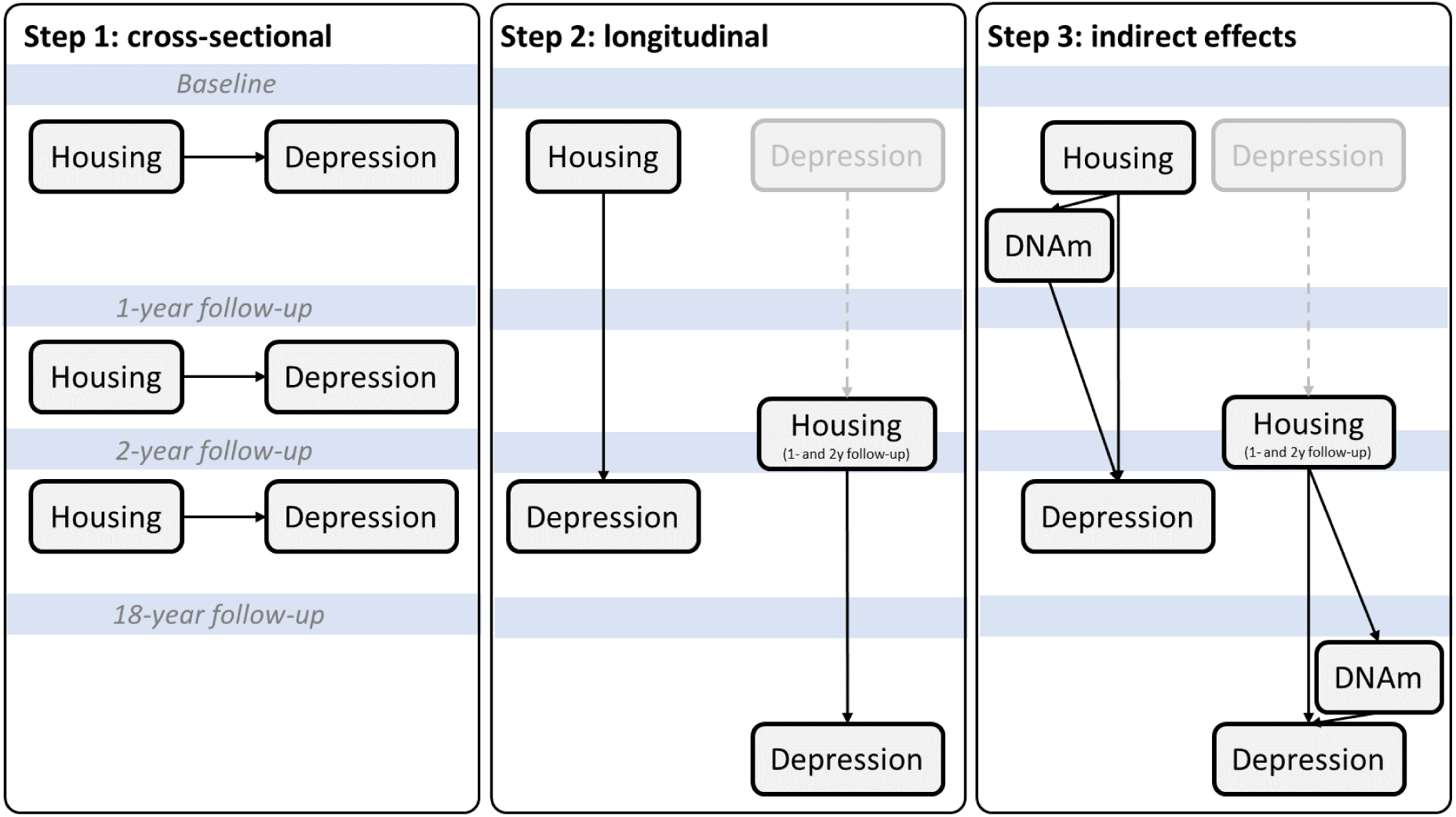
A) Step 1: Cross-sectional, B) Step 2: Longitudinal, and C) Step 3: DNAm indirect effects analyses overview. Depression at baseline (greyed-out box) was controlled for in all follow-up analyses. For a complete set of covariates, see SM Figure 1.

In step 2, we performed two longitudinal analyses. Specifically, we assessed whether poor housing quality (at study enrolment) associated with depressive symptoms 2 years later (mean age 30), controlling for SES risk factors, poor neighbourhood quality, housing instability and familial depressive history (model 4). Next, we assessed whether poor housing quality (at 1- and 2-year follow-ups) associated with depressive symptoms 18 years later (mean age 48), controlling for baseline depressive symptoms (at study enrolment), familial depressive history, housing instability, SES risk factors and poor neighbourhood quality (model 5). To minimize the risk for reverse causation, we controlled for baseline depressive symptoms at study enrolment and purposefully measured poor housing quality only after this baseline time point (i.e., only at 1- and 2-year follow-ups, but not at study enrolment).

In step 3, we tested for indirect effects in the associations between poor housing quality and depressive symptoms in both longitudinal models that were partly explained by DNAm. We applied high dimensional mediational analysis ^51,52^. In model 6, we investigated the extent to which DNAm measured at study enrolment explained associations between poor housing quality (at study enrolment) and depressive symptoms (at 2-year follow-up). In model 7, we tested whether DNAm measured at age 48 years partly explained the association between poor housing quality (at 1- and 2-year follow ups) and depressive symptoms (at 18-year follow-up).

Due to the large number of possible indirect effects (i.e., 485,000 CpGs), we used sure independence screening to prune CpGs down to a smaller set of potential intermediary variables ^53^ (see SM section 1.3). Cell type, smoking, batch, and age at DNAm assessment were regressed out of the dataset prior to conducting indirect effect analysis. Ethnicity was not regressed out as 98% of participants self-identified as white. To test for indirect effects, we used a sparse group lasso penalized model from the ‘regmed’ R package developed by Schaid & Sinnwell (^52^) in R Studio Statistical Software (v4.1.0; R Core Team, 2021) and applied to epigenetic data for the first time by Lussier and colleagues ^51^. This method was selected due to its increased power to detect indirect effects in comparison to structural equation modelling.

## CpG annotation

For CpG sites that showed evidence of an indirect effect, UCSC Genome Browser was used to identify the nearest gene (GRCh38/hg38; ^27^, EWAS atlas to identify CpG-trait associations (^28^; https://ngdc.cncb.ac.cn/ewas/atlas), and GWAS catalogue for gene-trait associations (^29^; https://www.ebi.ac.uk/gwas/) using default settings.

## Data Availability

Please note that the study website contains details of all the data that is available through a fully searchable data dictionary and variable search tool (http://www.bristol.ac.uk/alspac/researchers/our-data/). Access to ALSPAC data is through a system of managed open access (http://www.bristol.ac.uk/alspac/researchers/access/).

## Acknowledgements

We are extremely grateful to all the families who took part in this study, the midwives for their help in recruiting them, and the whole ALSPAC team, which includes interviewers, computer and laboratory technicians, clerical workers, research scientists, volunteers, managers, receptionists and nurses.

## References

1. Office for National Statistics. Is hybrid working here to stay? https://www.ons.gov.uk/employmentandlabourmarket/peopleinwork/employmentandemployeetypes/articles/ishybridworkingheretostay/2022-05-23 (2022).

2. Pevalin, D. J., Reeves, A., Baker, E. & Bentley, R. The impact of persistent poor housing conditions on mental health: A longitudinal population-based study. Prev. Med. 105, 304–310 (2017).

3. Singh, A., Daniel, L., Baker, E. & Bentley, R. Housing Disadvantage and Poor Mental Health: A Systematic Review. Am. J. Prev. Med. 57, 262–272 (2019).

4. Howden-Chapman, P. L., Chandola, T., Stafford, M. & Marmot, M. The effect of housing on the mental health of older people: the impact of lifetime housing history in Whitehall II. BMC Public Health 11, 682 (2011).

5. Lee, J. H. Housing quality determinants of depression and suicide ideation by age and gender. Hous. Stud. 39, 502–528 (2024).

6. Duncan, D. T. et al. The built environment and depressive symptoms among urban youth: A spatial regression study. Spat. Spatio-Temporal Epidemiol. 5, 11–25 (2013).

7. Lederbogen, F. et al. City living and urban upbringing affect neural social stress processing in humans. Nature 474, 498–501 (2011).

8. Peen, J., Schoevers, R. A., Beekman, A. T. & Dekker, J. The current status of urban-rural differences in psychiatric disorders. Acta Psychiatr. Scand. 121, 84–93 (2010).

9. Chapin, F. S. Some Housing Factors Related to Mental Hygiene. Am. J. Public Health Nations Health 41, 839–845 (1951).

10. Evans, G. W., Wells, N. M., Chan, H. Y. & Saltzman, H. Housing quality and mental health. J. Consult. Clin. Psychol. 68, 526–530 (2000).

11. Rollings, K. A., Wells, N. M., Evans, G. W., Bednarz, A. & Yang, Y. Housing and neighborhood physical quality: Children’s mental health and motivation. J. Environ. Psychol. 50, 17–23 (2017).

12. Dunn, J. R., Hayes, M. V., Hulchanski, J. D., Hwang, S. W. & Potvin, L. Housing as a Socio-Economic Determinant of Health: Findings of a National Needs, Gaps and Opportunities Assessment. Can. J. Public Health Rev. Can. Santé Publique 97, S12–S17 (2006).

13. Edwards, J. N., Edwards, P. K. & Booth, A. Housing Type, Stress, and Family Relations*. Soc. Forces 61, 241–257 (1982).

14. Adler, N. E. & Snibbe, A. C. The Role of Psychosocial Processes in Explaining the Gradient between Socioeconomic Status and Health. Curr. Dir. Psychol. Sci. 12, 119–123 (2003).

15. Wagstaff, A. & Watanabe, N. What difference does the choice of SES make in health inequality measurement? Health Econ. 12, 885–890 (2003).

16. Dohrenwend, B. P. et al. Socioeconomic status and psychiatric disorders: the causation-selection issue. Science 255, 946–952 (1992).

17. Colodro-Conde, L. et al. Association Between Population Density and Genetic Risk for Schizophrenia. JAMA Psychiatry 75, 901–910 (2018).

18. Dunn, E. C. The Role of Neighborhood Social Characteristics on the Epigenome—Why the Lack of Investigations? (2020) doi:10.1001/jamanetworkopen.2020.6111.

19. Dunn, E. C. et al. Research review: gene-environment interaction research in youth depression - a systematic review with recommendations for future research. J. Child Psychol. Psychiatry 52, 1223–1238 (2011).

20. Lesch, K. P. Gene–environment interaction and the genetics of depression. J. Psychiatry Neurosci. 29, 174–184 (2004).

21. Siegfried, Z. & Simon, I. DNA methylation and gene expression. Wiley Interdiscip. Rev. Syst. Biol. Med. 2, 362–371 (2010).

22. Jeong, A. et al. Residential greenness-related DNA methylation changes. Environ. Int. 158, 106945 (2022).

23. Lussier, A. A. et al. Association between the timing of childhood adversity and epigenetic patterns across childhood and adolescence: findings from the Avon Longitudinal Study of Parents and Children (ALSPAC) prospective cohort. *Lancet Child Adolesc*. Health 7, 532–543 (2023).

24. Marini, S. et al. Adversity exposure during sensitive periods predicts accelerated epigenetic aging in children. Psychoneuroendocrinology 113, 104484 (2020).

25. Barker, E. D., Walton, E. & Cecil, C. A. M. Annual Research Review: DNA methylation as a mediator in the association between risk exposure and child and adolescent psychopathology. J. Child Psychol. Psychiatry 59, 303–322 (2018).

26. Howden-Chapman, P. et al. Review of the Impact of Housing Quality on Inequalities in Health and Well-Being. Annu. Rev. Public Health 44, 233–254 (2023).

27. Kent, W. J. et al. The human genome browser at UCSC. Genome Res. 12, 996–1006 (2002).

28. Li, M. et al. EWAS Atlas: a curated knowledgebase of epigenome-wide association studies. Nucleic Acids Res. 47, D983–D988 (2019).

29. Sollis, E. et al. The NHGRI-EBI GWAS Catalog: knowledgebase and deposition resource. Nucleic Acids Res. 51, D977–D985 (2023).

30. Cecil, C. A. M., Neumann, A. & Walton, E. Epigenetics applied to child and adolescent mental health: Progress, challenges and opportunities. JCPP Adv. 3, e12133, jcv2.12133 (2023).

31. Wendt, F. R. et al. Multivariate genome-wide analysis of education, socioeconomic status and brain phenome. *Nat*. Hum. Behav. 5, 482–496 (2021).

32. Wang, H. et al. Genome-wide interaction analysis of pathological hallmarks in Alzheimer’s disease. Neurobiol. Aging 93, 61–68 (2020).

33. Shrine, N. et al. New genetic signals for lung function highlight pathways and chronic obstructive pulmonary disease associations across multiple ancestries. Nat. Genet. 51, 481–493 (2019).

34. Chang, X. et al. Genetic associations with healthy ageing among Chinese adults. Npj Aging 8, 6 (2022).

35. Jokela, M. & Laakasuo, M. Obesity as a causal risk factor for depression: Systematic review and meta-analysis of Mendelian Randomization studies and implications for population mental health. J. Psychiatr. Res. 163, 86–92 (2023).

36. Wang, C. et al. Short- and intermediate-term exposure to ambient fine particulate elements and leukocyte epigenome-wide DNA methylation in older men: the Normative Aging Study. Environ. Int. 158, 106955 (2022).

37. Leventhal, T. & Brooks-Gunn, J. Moving to Opportunity: an Experimental Study of Neighborhood Effects on Mental Health. Am. J. Public Health 93, 1576–1582 (2003).

38. Boyd, A. et al. Cohort Profile: The ‘Children of the 90s’—the index offspring of the Avon Longitudinal Study of Parents and Children. Int. J. Epidemiol. 42, 111–127 (2013).

39. Fraser, A. et al. Cohort Profile: the Avon Longitudinal Study of Parents and Children: ALSPAC mothers cohort. Int. J. Epidemiol. 42, 97–110 (2013).

40. Major-Smith, D. et al. The Avon Longitudinal Study of Parents and Children (ALSPAC): a 2022 update on the enrolled sample of mothers and the associated baseline data. Wellcome Open Res. 7, 283 (2023).

41. Relton, C. L. et al. Data Resource Profile: Accessible Resource for Integrated Epigenomic Studies (ARIES). Int. J. Epidemiol. 44, 1181–1190 (2015).

42. Department for Levelling Up, Housing and Communities. A decent home: definition and guidance. GOV.UK https://www.gov.uk/government/publications/a-decent-home-definition-and-guidance (2006).

43. Crown, S. & Crisp, A. H. A Short Clinical Diagnostic Self-rating Scale for Psychoneurotic Patients: The Middlesex Hospital Questionnaire (M.H.Q.). Br. J. Psychiatry 112, 917–923 (1966).

44. Newnham, E. A., Harwood, K. E. & Page, A. C. Evaluating the clinical significance of responses by psychiatric inpatients to the mental health subscales of the SF-36. J. Affect. Disord. 98, 91–97 (2007).

45. Elliott, T. E., Renier, C. M. & Palcher, J. A. Chronic pain, depression, and quality of life: correlations and predictive value of the SF-36. Pain Med. Malden Mass 4, 331–339 (2003).

46. Burgess, P. M., Mazzocco, L. & Campbell, I. M. Discriminant validity of the Crown-Crisp Experiential Index. Br. J. Med. Psychol. 60, 61–69 (1987).

47. Fukuhara, S., Ware, J. E., Kosinski, M., Wada, S. & Gandek, B. Psychometric and clinical tests of validity of the Japanese SF-36 Health Survey. J. Clin. Epidemiol. 51, 1045–1053 (1998).

48. Min, J. L., Hemani, G., Davey Smith, G., Relton, C. & Suderman, M. Meffil: efficient normalization and analysis of very large DNA methylation datasets. Bioinforma. Oxf. Engl. 34, 3983–3989 (2018).

49. Price, E. M. et al. Additional annotation enhances potential for biologically-relevant analysis of the Illumina Infinium HumanMethylation450 BeadChip array. Epigenetics Chromatin 6, 4 (2013).

50. Reinius, L. E. et al. Differential DNA methylation in purified human blood cells: implications for cell lineage and studies on disease susceptibility. PloS One 7, e41361 (2012).

51. Lussier, A. A. et al. DNA methylation mediates the link between adversity and depressive symptoms. Nat. Ment. Health (2024).

52. Schaid, D. J. & Sinnwell, J. P. Penalized Models for Analysis of Multiple Mediators. Genet. Epidemiol. 44, 408–424 (2020).

53. Fan, J. & Lv, J. Sure independence screening for ultrahigh dimensional feature space. J. R. Stat. Soc. Ser. B Stat. Methodol. 70, 849–911 (2008).

