## Supplementary Materials for "Home and Epigenome: DNA Methylation as a Link Between Poor Housing Quality and Depressive Symptoms"

**Home and Epigenome: Indirect effect of DNA methylation between poor housing quality and depression association**

**Affiliations:**

Supplementary Materials

**1 Methods**

**1.1 Missing Data Imputations**

Mothers who had completed questionnaire items for their housing circumstances at study enrolment or, 1- or 2-year follow-ups were selected, leaving up to 9,566 mothers available for analysis. Missing data for the exposure, covariates, and outcomes were imputed. Imputation was conducted using the R MICE package ^1^. The exposure (poor housing quality) and all covariates were used as predictors in a predictor matrix to impute missing data (in poor housing quality, all covariates and outcomes. Covariates included SES risk factors, familial depressive history, poor neighbourhood quality, age at depressive symptoms measurement, smoking, ethnicity, age at DNAm, age at housing and housing instability. Poor housing quality scores were calculated if more than 25% of the variables that comprise each score were available (prior to imputation). However, if a person had more than 25% missingness, an NA score was assigned and then later imputed using passive imputation ^2^. Passive imputation refers to where input variables (e.g., housing decorations) were imputed first using predictive mean matching before calculating row means (e.g., poor housing quality), increasing the accuracy and power of imputations ^2^.

Imputation was performed using 60 iterations and 30 imputed datasets. The predictor matrix was specified using quickpred(), with a minimum correlation set to 0.05, striking a balance between having sufficient variable breadth yet avoiding weak predictors. The predictor matrix was modified so outcomes (depressive symptom scores) would be imputed but not used as predictors.

**1.2 Poor Neighbourhood Quality**

Neighbourhood quality was self-reported at study enrolment. Participants were asked if their neighbourhood was ‘lively’, ‘friendly’, ‘noisy’, ‘clean’, ‘attractive’, ‘polluted/dirty’. Participants had to respond to each of these characteristics with either, ‘Yes usually’, ‘Yes sometimes’, or ‘No not at all’. Neighbourhood quality was self-reported at 2-year follow-up. Participants were asked, ‘Here is a list of things that can be a problem in people’s homes or in the neighbourhood. How much of a problem are the following for you and your family?’, followed by ‘Badly fitted windows and doors’, ‘Poor ventilation’, ‘Noise travelling between the rooms of your home’, ‘Noise from other homes’, ‘Noise from outside in the street’, ‘Rubbish or litter dumped around your neighbourhood’, ‘Dog dirt on pavements/walkways’, ‘Worry about vandalism’, ‘Worry about burglars’, ‘Worry about muggings or attacks’, ‘Disturbance from teenagers or youths’. Participants had to respond to each of the items on this list with, ‘Serious problem’, ‘Minor problem’, ‘Not a problem’ or ‘No opinion’.

**1.3 Statistical Analysis and CpG Pruning**

**SM Figure 1.** A) Step 1: Cross-sectional, B) Step 2: Longitudinal, and C) Step 3: DNAm analyses overview. The timeline along the x axis indicates timepoints variables were measured at. The type of variable (i.e., exposure, covariate, intermediary variable, or outcome) is categorised along the y axis. Models are indicated by colour. *Familial depressive history is used in all models.

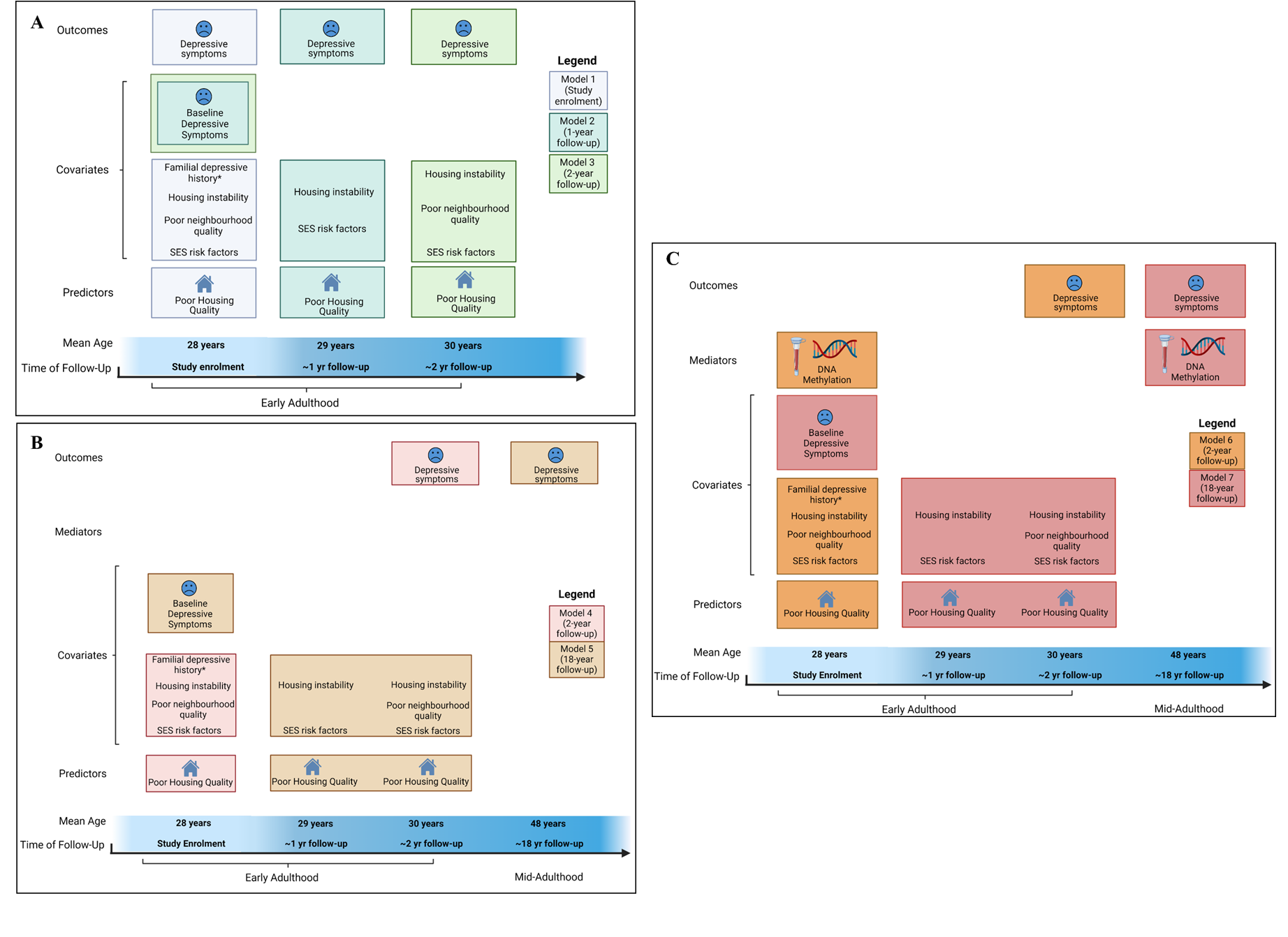

DNAm data has a large number of potential intermediary variables (i.e. 485,000 CpGs). Hence, to reduce this number, a high dimensional mediational analysis method was followed using sure independence screening (SIS; ^3^).

In SIS, DNAm intermediary variables are reduced to a more viable number (*q*),

*q* = (n − 3 − 1)/2

where n equals the sample size, reducing the likelihood of detecting false positives. In model 6, *q* was equal to 425 CpGs, and in model 7, *q* was equal to 364 CpGs. This equation maximizes the number of intermediary variables tested, while remaining below the number of parameters to be estimated given the sample size.

CpGs were reduced to the prespecified q, by selecting the top sites based on their marginal correlation with poor housing quality and depression ^4,5^:

*| cor(x, m_j_) ⋅ cor (m_j_, y) |*

where *x* is poor housing quality, *m_j_* is a given CpG site, and *y* is depressive symptoms. This step assumes CpGs showing stronger correlations with both the exposure (poor housing quality) and outcome (depressive symptoms) are more likely to be intermediary variables than CpGs with lower or no correlation with the exposure and outcome ^3^.

The top 425 and 364 potential intermediary sites for models 6 and 7, respectively, were entered into a structural equation model (SEM) using the regmed.grid function in the R package ‘regmed’. This algorithm identified CpG sites with the strongest indirect effect. Iterating through ten values for the shrinkage parameter of lambda, from one (the most conservative penalty) to zero (no penalty) in 0.1 increments, help identify the number of model parameters with the lowest Bayesian Information Criterion (BIC), reducing overfitting.

**2 Results**

**2.1 Descriptive results for all sub-samples.**

**Table S1.**

*Study descriptives for all models investigated in this study including both full sample and DNAm subsamples (imputed data). Statistical test results showing differences between full sample and DNAm subsamples (imputed data).
 ^1^ Depressive symptoms measured at enrolment, and at ~1 and ~2-year follow-up used the CCEI. Depressive symptoms measured at ~18-year follow-up used the SF-36 mental health scale.
^2^ Ethnicity (e.g., black/Caribbean, black/African, black/other, Indian, Pakistani, Bangladeshi, Chinese, any other ethnic group) was self-reported at study enrolment.
SD = standard deviation; SES = socioeconomic status.*

|  | **Cross-sectional** | | | **Longitudinal** | |  | | **DNAm Indirect Effects** | |  |
| --- | --- | --- | --- | --- | --- | --- | --- | --- | --- | --- |
|  | Enrolment  (n = 9,566) | 1-yr follow-up  (n = 9,566) | 2-yr follow-up  (n = 9,566) | ~2yr | ~18yr follow-up (n = 9,566) | ~2yr |  | | ~18yr |  |
|  |  |  |  | follow-up (n = 9,566) |  | follow-up (n = 871) | Full-vs-subsample difference | | follow-up  (n = 731) | Full-vs-subsample difference |
| **Poor** **Housing Quality** | | | | | | |  | |  |  |
| Mean (SD) | 3.44 (0.34) | 1.9 (0.22) | 2.97 (0.37) | 3.44 (0.34) | 2.44 (0.21) | 3.38 (0.35) | p=0.0009 | | 2.45 (0.21) | p=0.179 |
| Min, Max | 1.21, 4.81 | 1.01, 3.47 | 1.05, 4.27 | 1.21, 4.81 | 1.47, 3.22 | 1.21, 4.43 |  | | 1.49, 3.20 |  |
| **SES Risk Factors** | | | | | | | | | |  |
| Mean (SD) | 1.63 (0.88) | 2.02 (0.93) | 1.96 (0.94) | 1.63 (0.88) | 1.99 (0.86) | 1.49 (0.84) | p< 0.0001 | | 1.81 (0.83) | p<0.0001 |
| Min, Max | 0.60, 5.60 | 0.80, 6.20 | 0.80, 6.80 | 0.60, 5.60 | 0.80, 6.40 | 0.60, 4.80 |  | | 0.80, 4.90 |  |
| **Familial Depressive History** | | | | | | | | | |  |
| Both parents | 5.27% | | | 5.27% | | 1.72% | p=0.247 | | 1.64% | p=0.304 |
| One parent | 20.64% | | | 20.64% | | 22.38% |  | | 23.66% |  |
| None | 74.10% | | | 74.10% | | 75.89% |  | | 74.69% |  |
| **Housing Instability** | | | | | | | | | |  |
| Mean (SD) | 1.65 (1.75) | 1.40 (1.71) | 0.23 (0.55) | 1.65 (1.75) | 0.82 (0.94) | 1.56 (1.54) | p=0.142 | | 0.72 (0.83) | p=0.002 |
| Min, Max | 0.00, 28.00 | 0.00, 22.00 | 0.00, 8.00 | 0.00, 28.00 | 0.00, 11.50 | 0.00, 11.00 |  | | 0.00, 7.00 |  |
| **Poor Neighbourhood Quality** | | | | | | | | | |  |
| Mean | 4.74 (2.19) | NA | 3.89 (3.52) | 4.74 (2.19) | 3.89 (3.52) | NA |  | | 3.80 (2.99) | p=0.465 |
| Min, Max | 1.00, 13.00 |  | 0.00, 22.00 | 1.00, 13.00 | 0.00, 22.00 |  | NA | | 0.00, 18.00 |  |
| **Depressive Symptoms at baseline (covariate)** | | | | | | | | | |  |
| Mean | NA ^1^ | 4.30 (3.02) | | NA | 4.30 (3.02) | 4.04 (2.79) |  | | 4.01 (2.73) | p=0.006 |
| Min, Max |  | 0.00, 16.00 | |  | 0.00, 16.00 | 0.00, 16.00 | NA | | 0.00, 16.00 |  |
| **Depressive Symptoms at follow-up** **^1^ (outcome)** | | | | | | | | | |  |
| Mean | 4.30 (3.02) | 3.30 (2.97) | 2.91 (2.68) | 2.91 (2.68) | 14.70 (4.38) | 2.84 (2.55) | p=0.459 | | 14.24 (4.23) | p=0.003 |
| Min, Max | 0.00, 16.00 | 0.00, 16.00 | 0.00, 14.00 | 0.00, 14.00 | 3.00, 25.00 | 0.00, 14.00 |  | | 3.00, 25.00 |  |
| **Age at Poor Housing Quality** | | | | | | | | | |  |
| Mean | 28.37 (4.68) | 29.46 (4.68) | 29.99 (4.73) | 28.37 (4.68) | 29.73 (4.71) | 29.27 (4.26) | p<0.0001 | | 30.87 (4.24) | p<0.0001 |
| Min, Max | 15.00, 44.00 | 16.00, 45.00 | 15.00, 63.00 | 15.00, 44.00 | 15.00, 63.00 | 17.00, 42.00 |  | | 19.00, 48.00 |  |
| **Age at Depressive Symptoms at follow-up** | | | | | | | | | |  |
| Mean | 28.36 (4.67) | 29.46 (4.68) | 29.99 (4.73) | 29.99 (4.73) | 47.71 (4.72) | 30.89 (4.34) | p<0.0001 | | 48.79 (4.21) | p<0.0001 |
| Min, Max | 15.00, 44.00 | 16.00, 45.00 | 15.00, 63.00 | 15.00, 63.00 | 34.00, 63.00 | 19.00, 48.00 |  | | 34.00, 61.00 |  |
| **Ethnicity ^2^** | | | | | | | | | |  |
| White | 98.12% | | | 98.12% | | 98.97% | p=0.072 | | 98.77% | p=0.170 |
| Non-White | 1.88% | | | 1.88% | | 1.03% |  | | 1.23% |  |
| **Smoking at enrolment** | | | | | | | | | |  |
| Never | 51.94% | | | 51.94% | NA | 62.57% | p<0.0001 | | NA |  |
| Until pregnancy | 29.70% | | | 29.70% |  | 26.63% |  | |  | NA |
| During pregnancy | 18.35% | | | 18.35% |  | 10.79% |  | |  |  |
| **Smoking at ~18y follow-up** | | | | | | | | | |  |
| 0 per day | NA | | | NA | 53.38% | NA |  | | 63.34% | p<0.0001 |
| 1-15 per day |  |  |  |  | 29.99% |  | NA | | 24.35% |  |
| 20+ per day |  |  |  |  | 16.63% |  |  | | 12.31% |  |

**2.2 Directions of methylation patterns**

**SM Figure 2:** Model 6 Direction of associations between poor housing quality and DNAm (left column), and DNAm and depressive symptoms (right column).

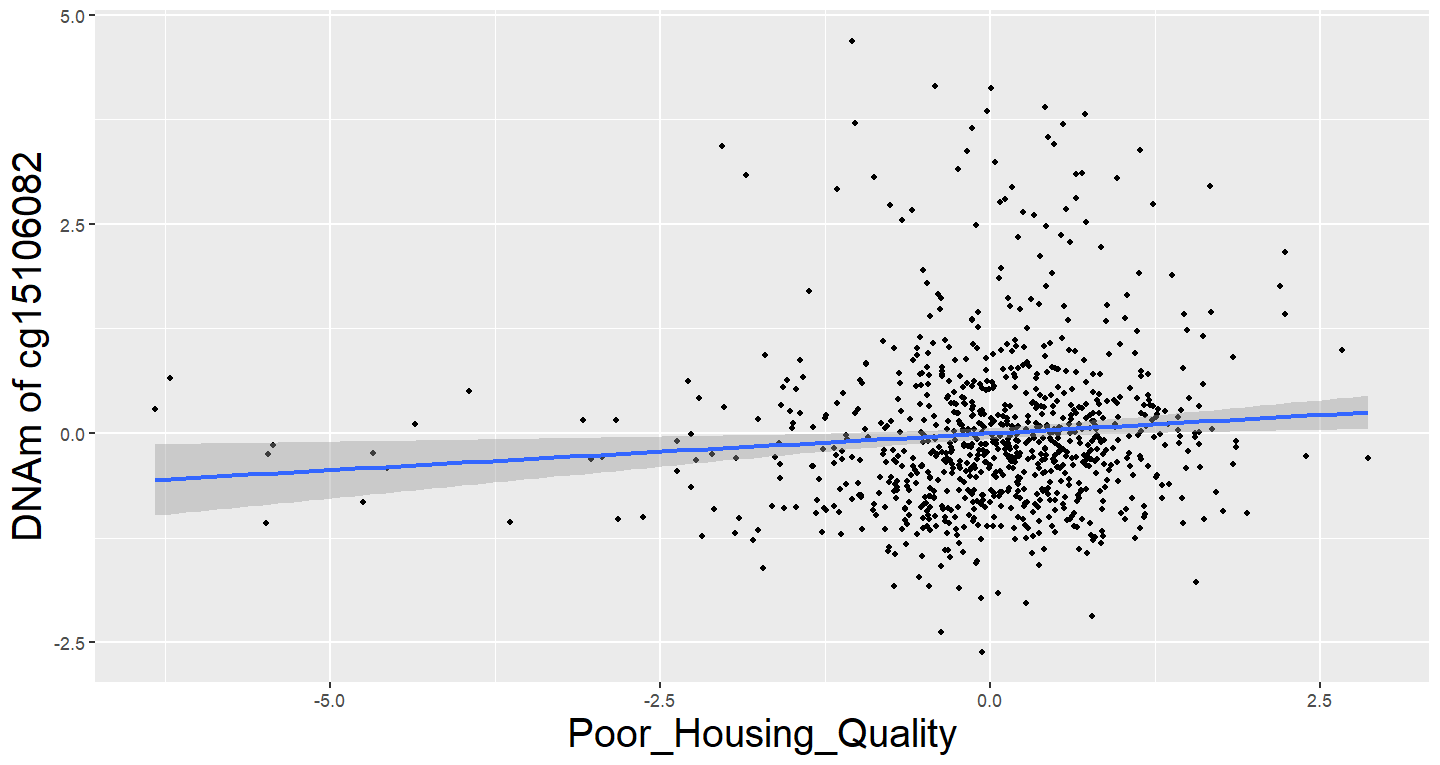

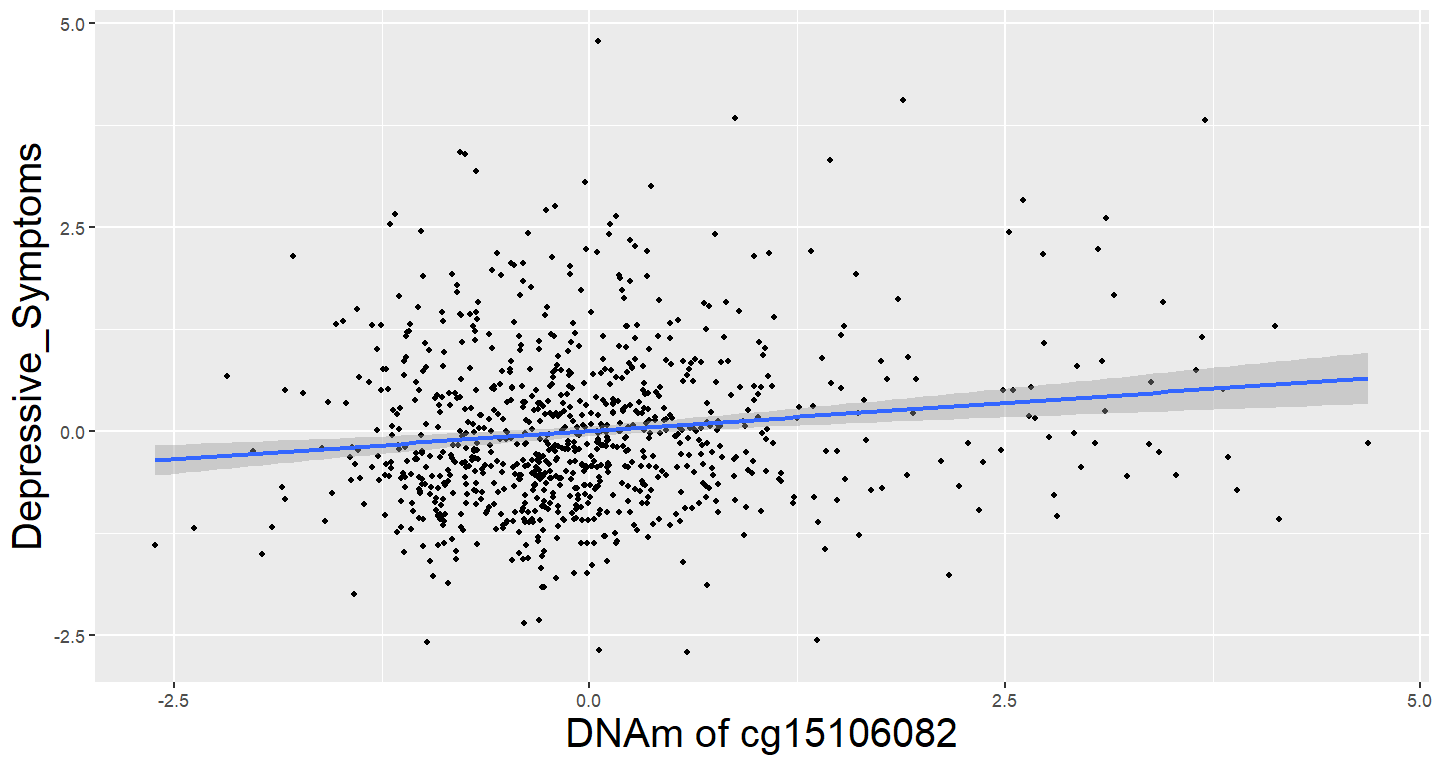

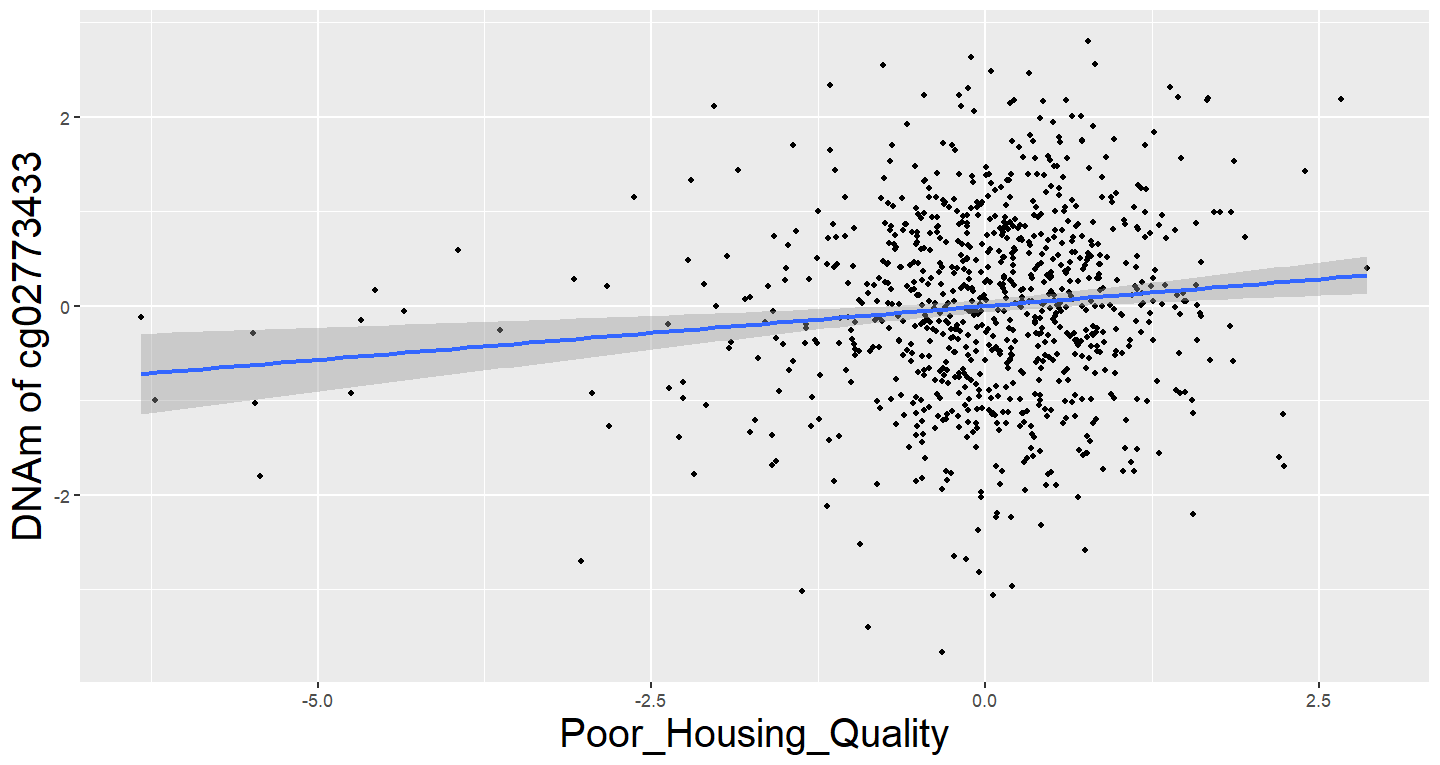

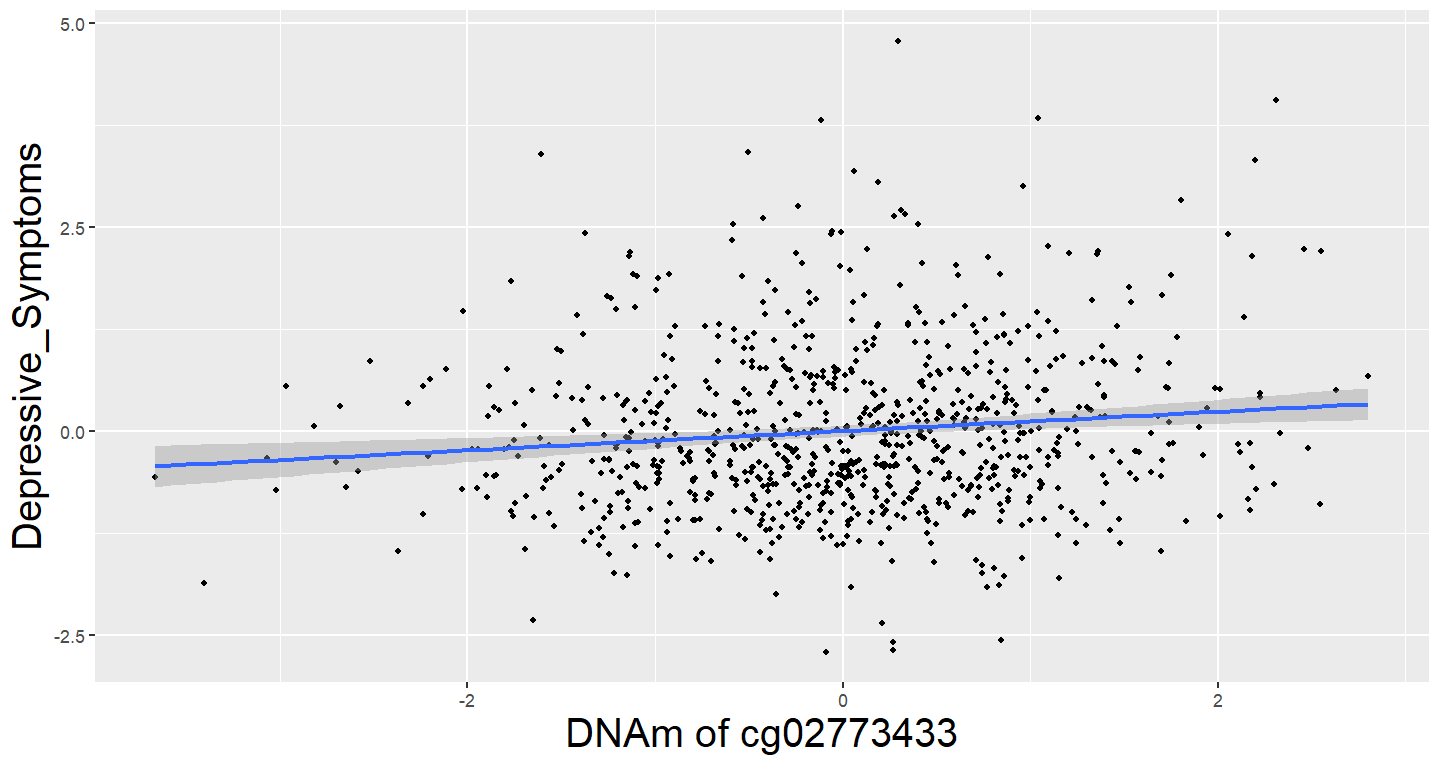

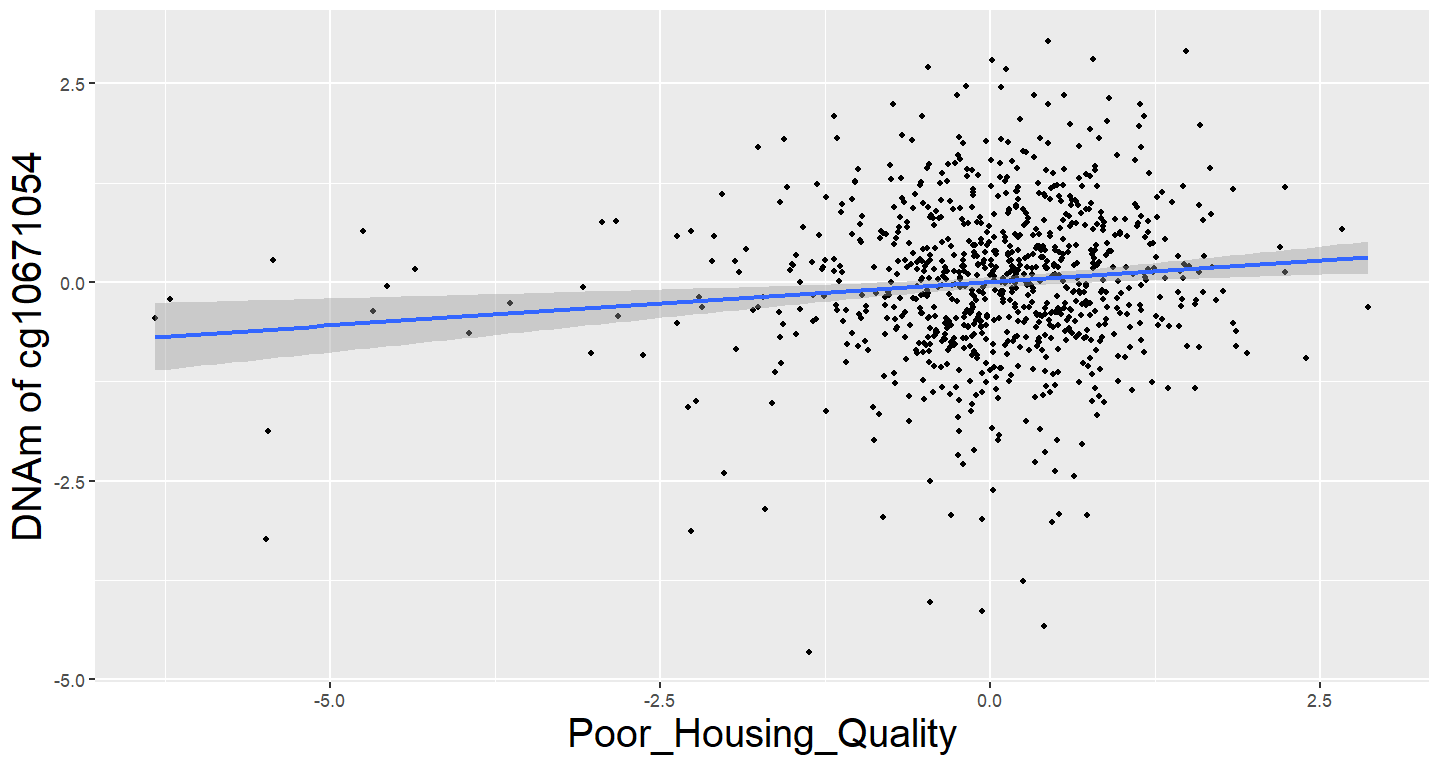

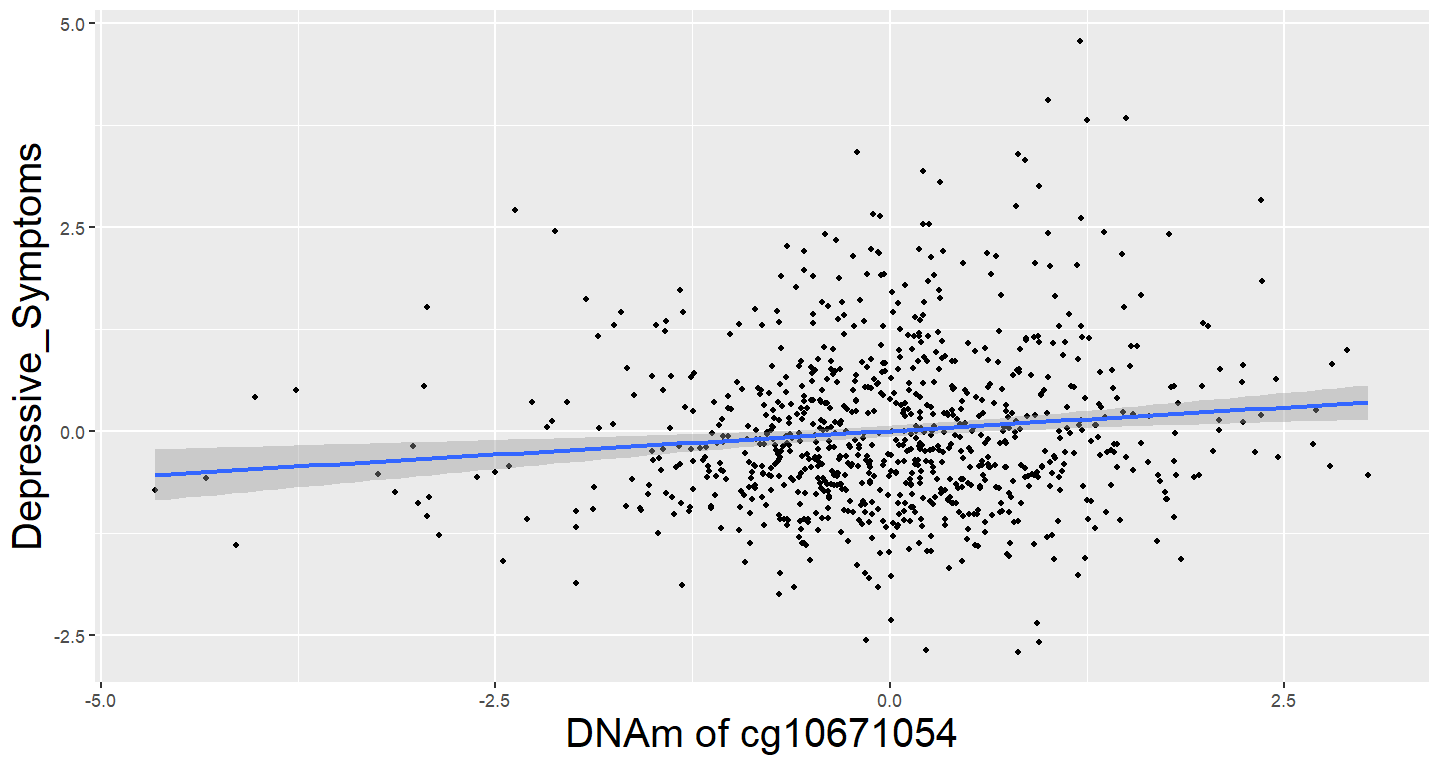

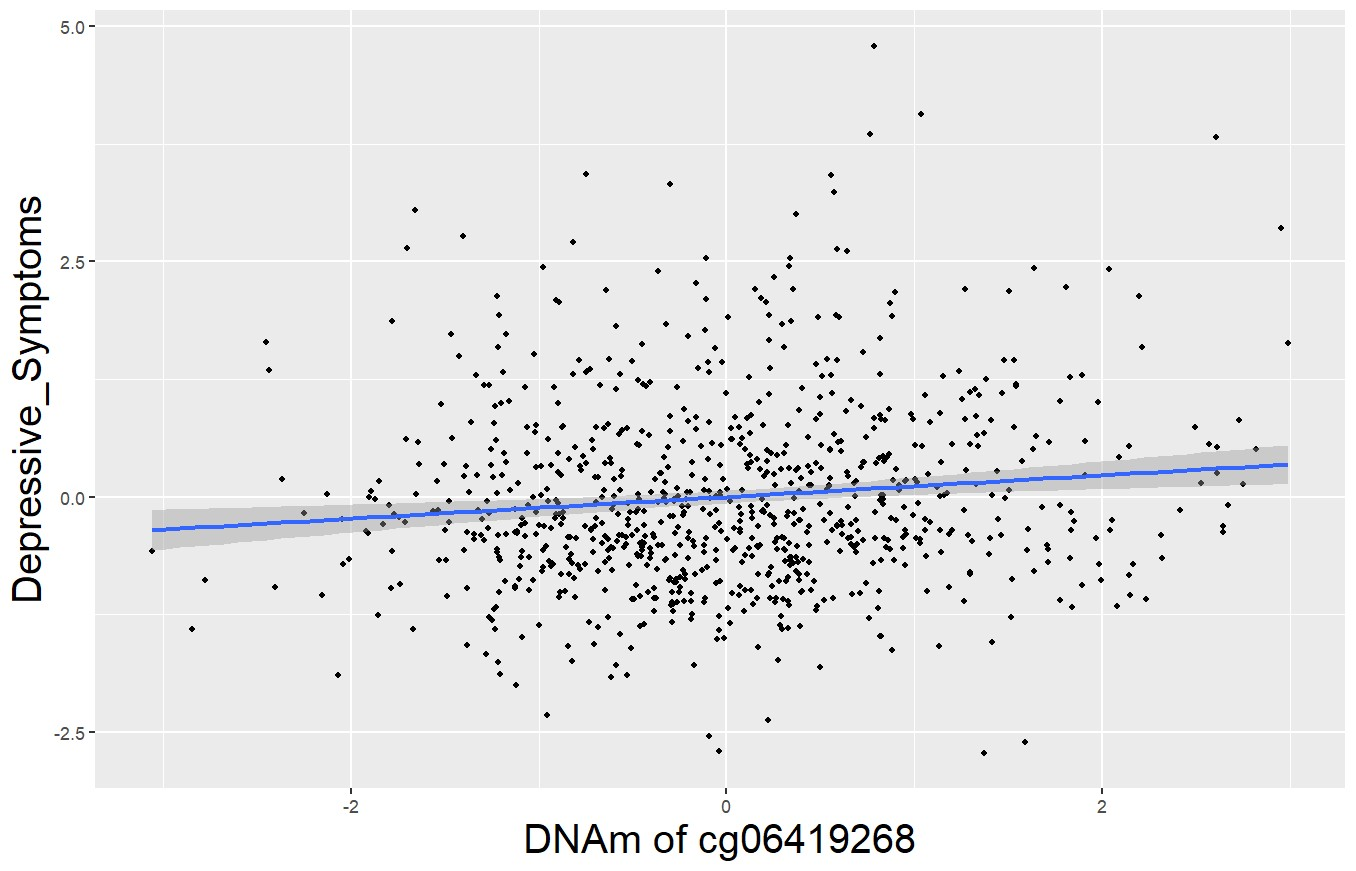

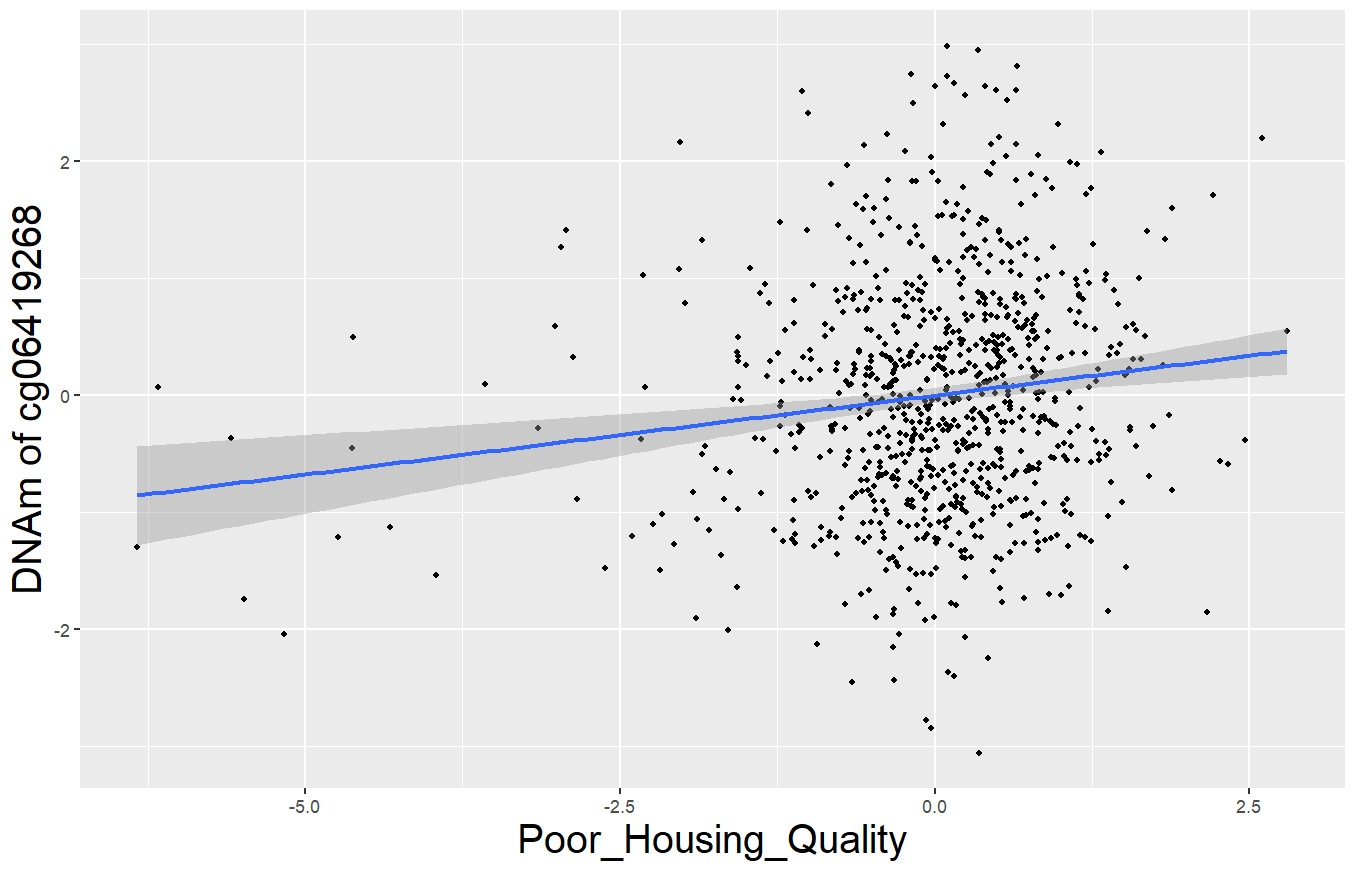

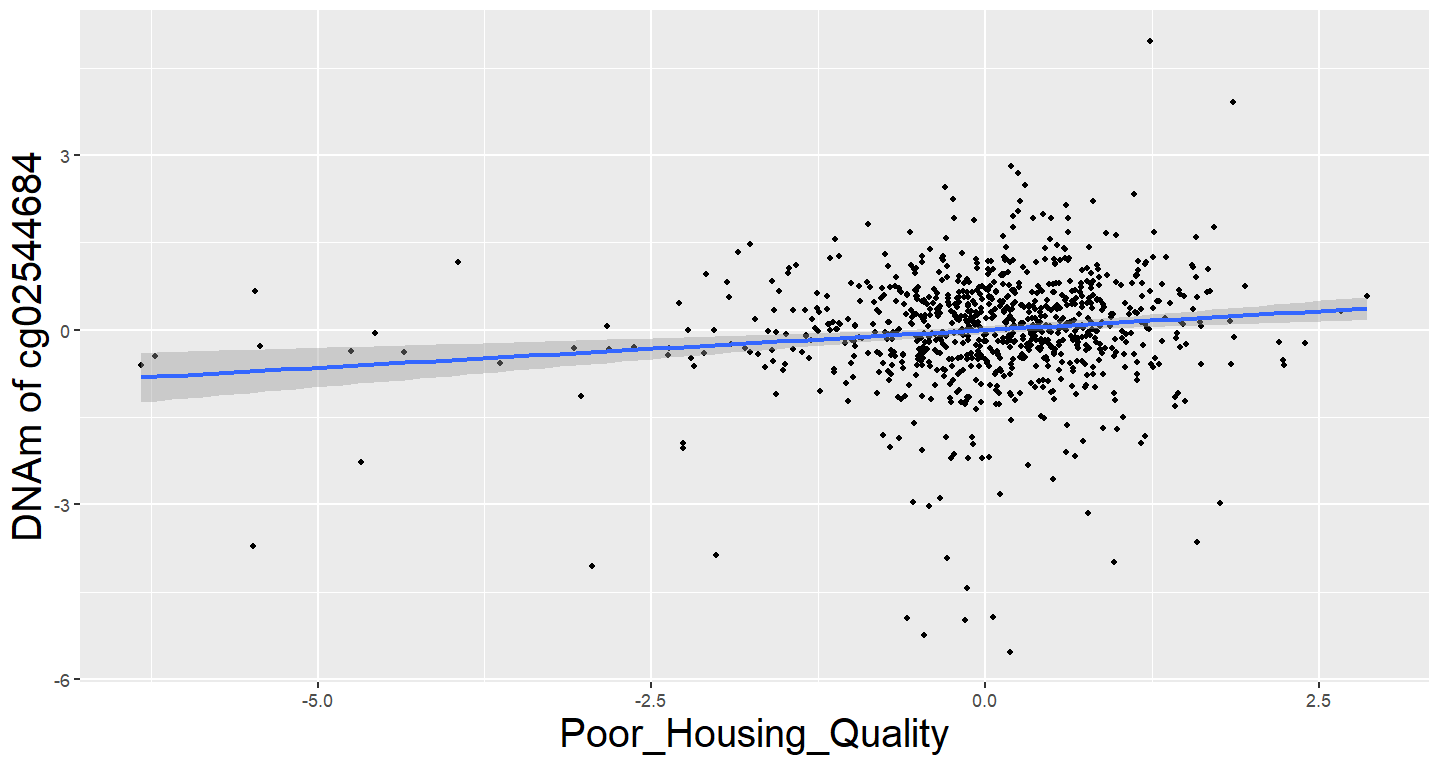

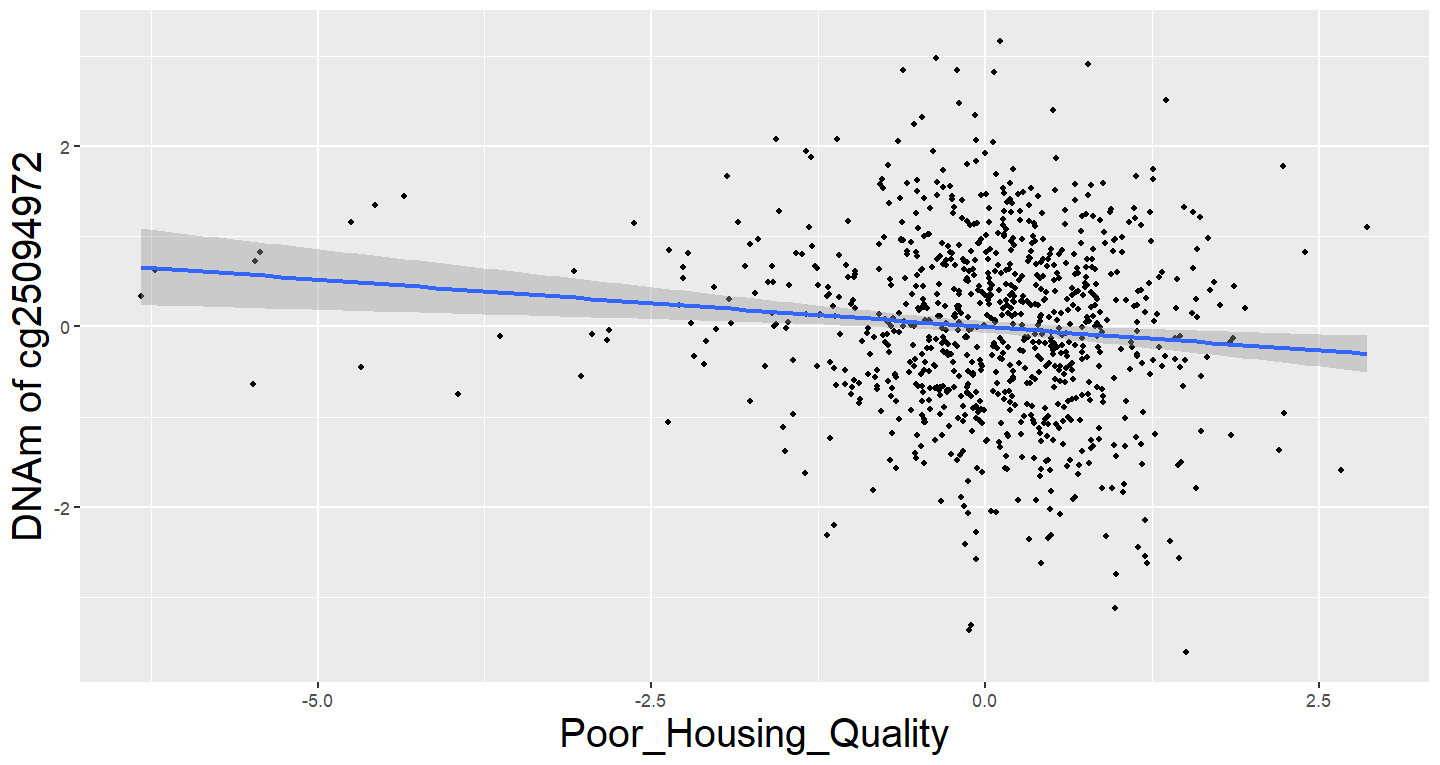

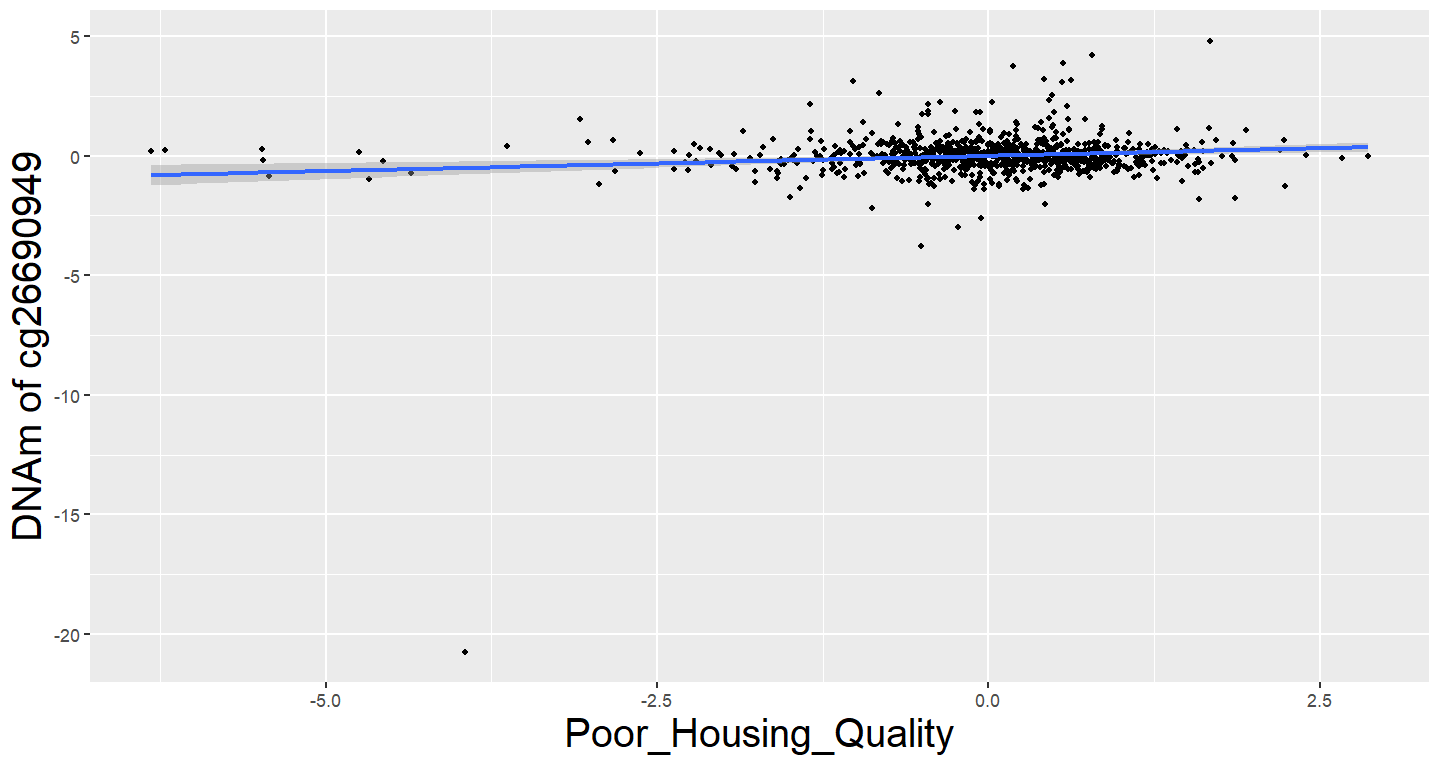

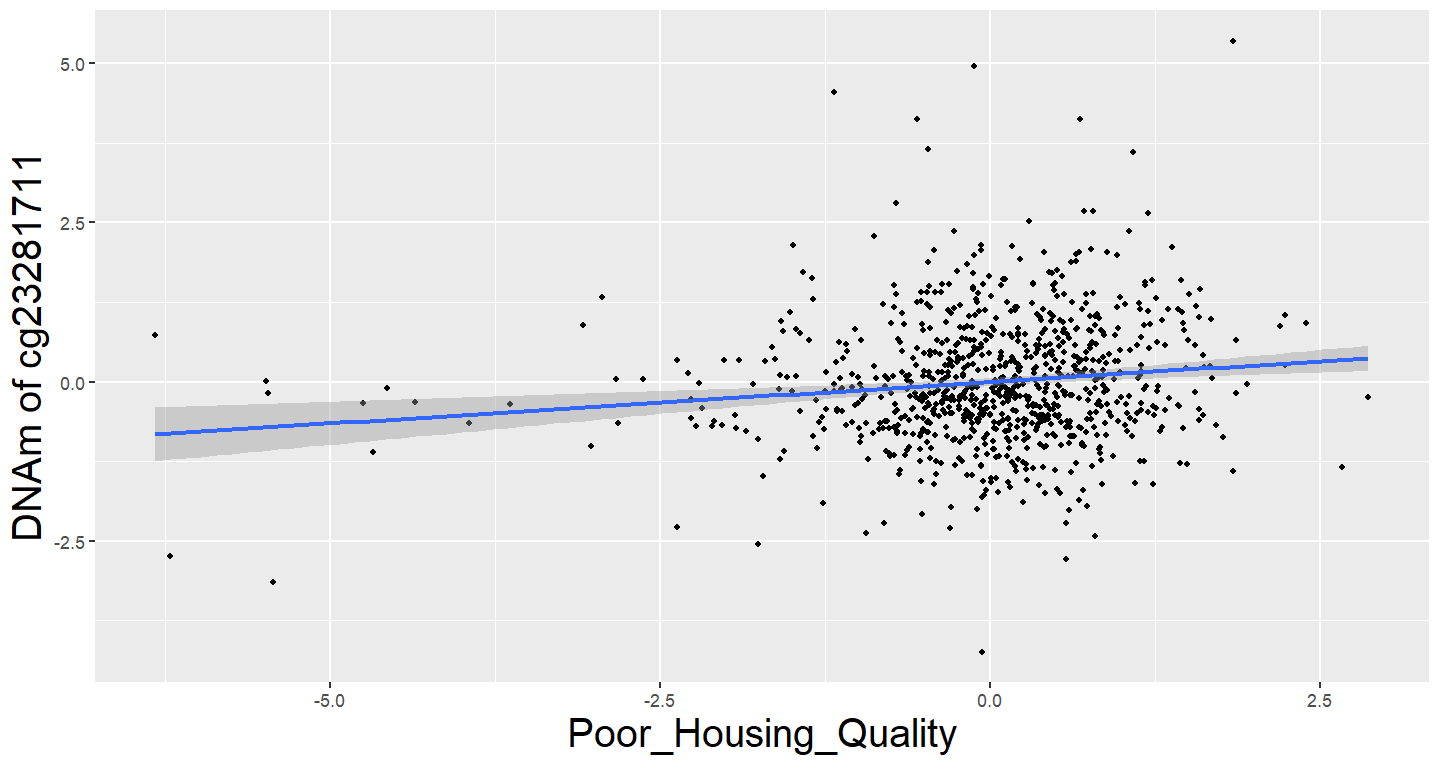

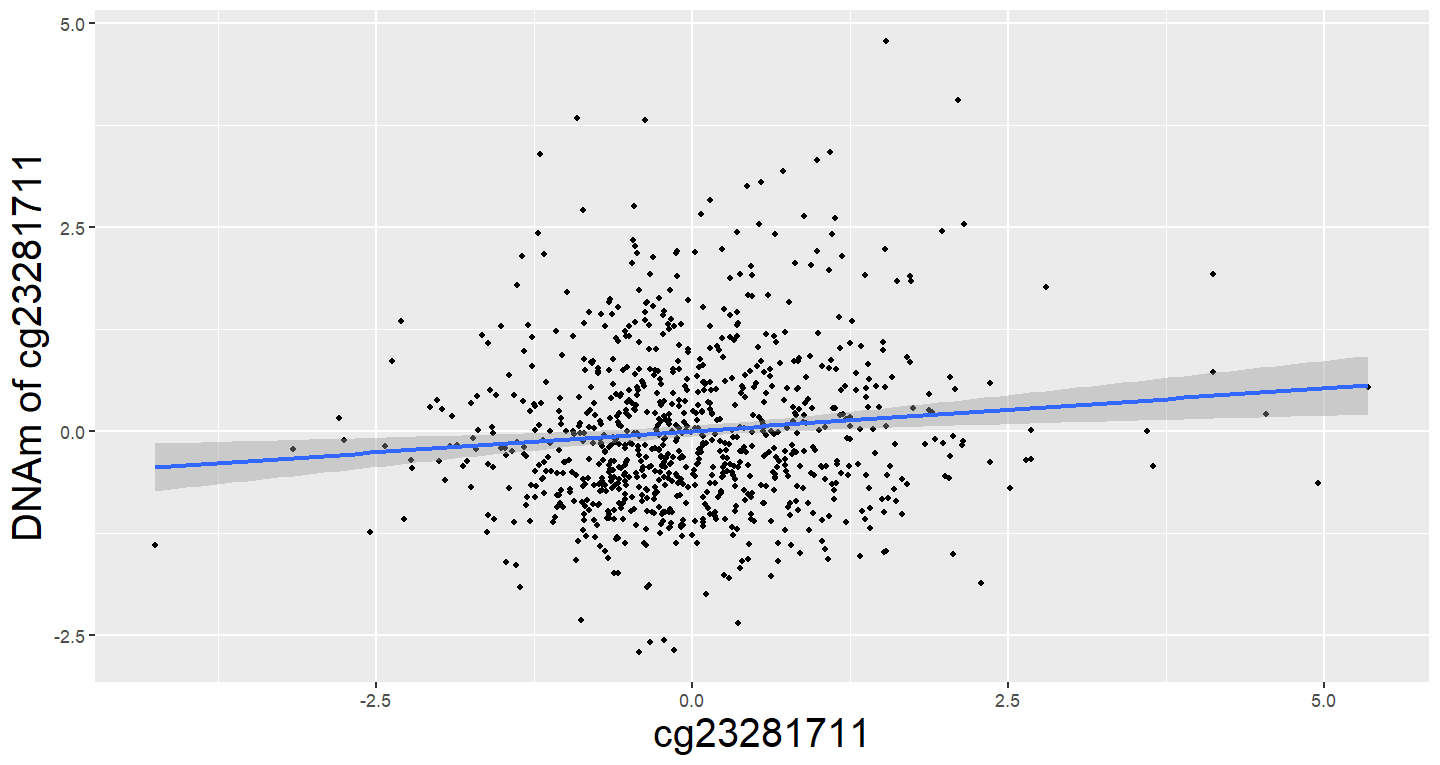

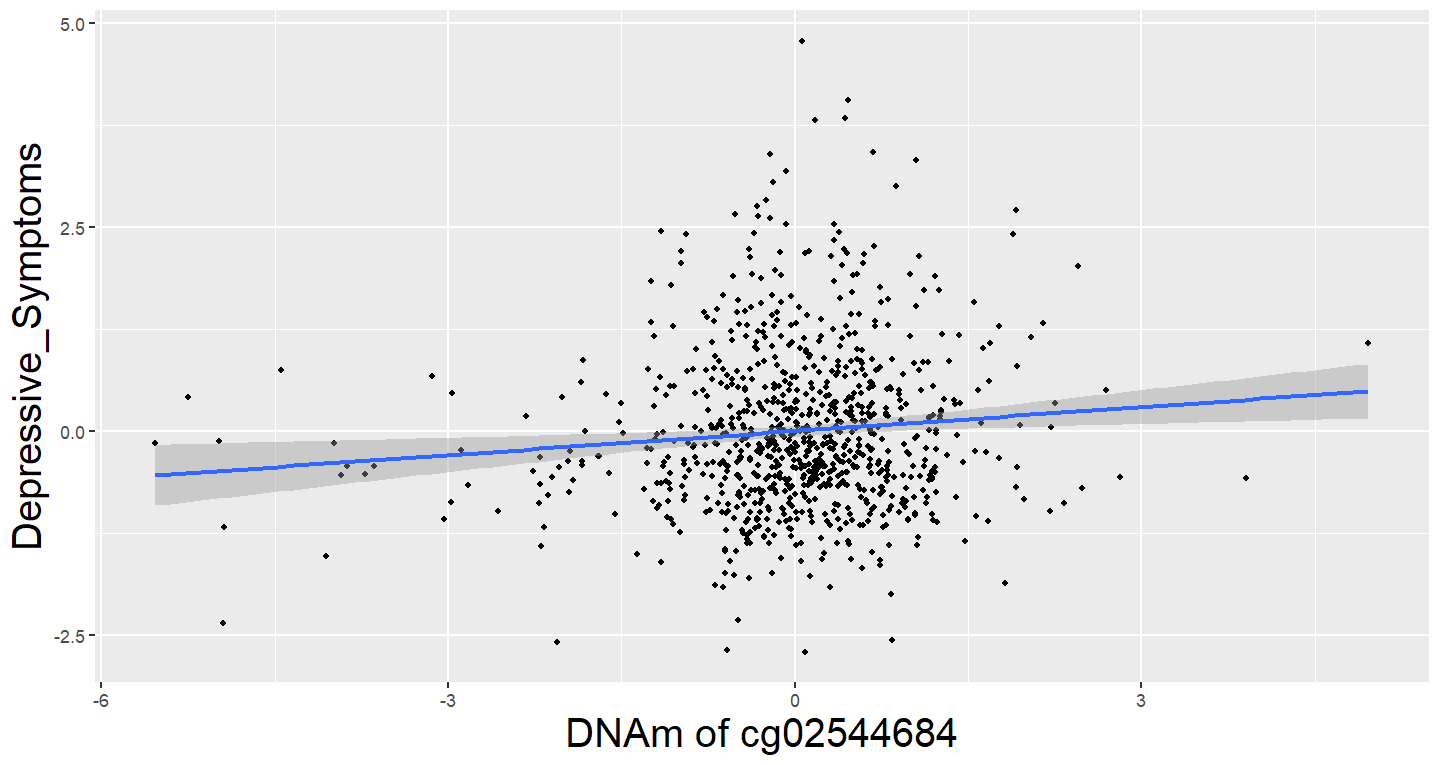

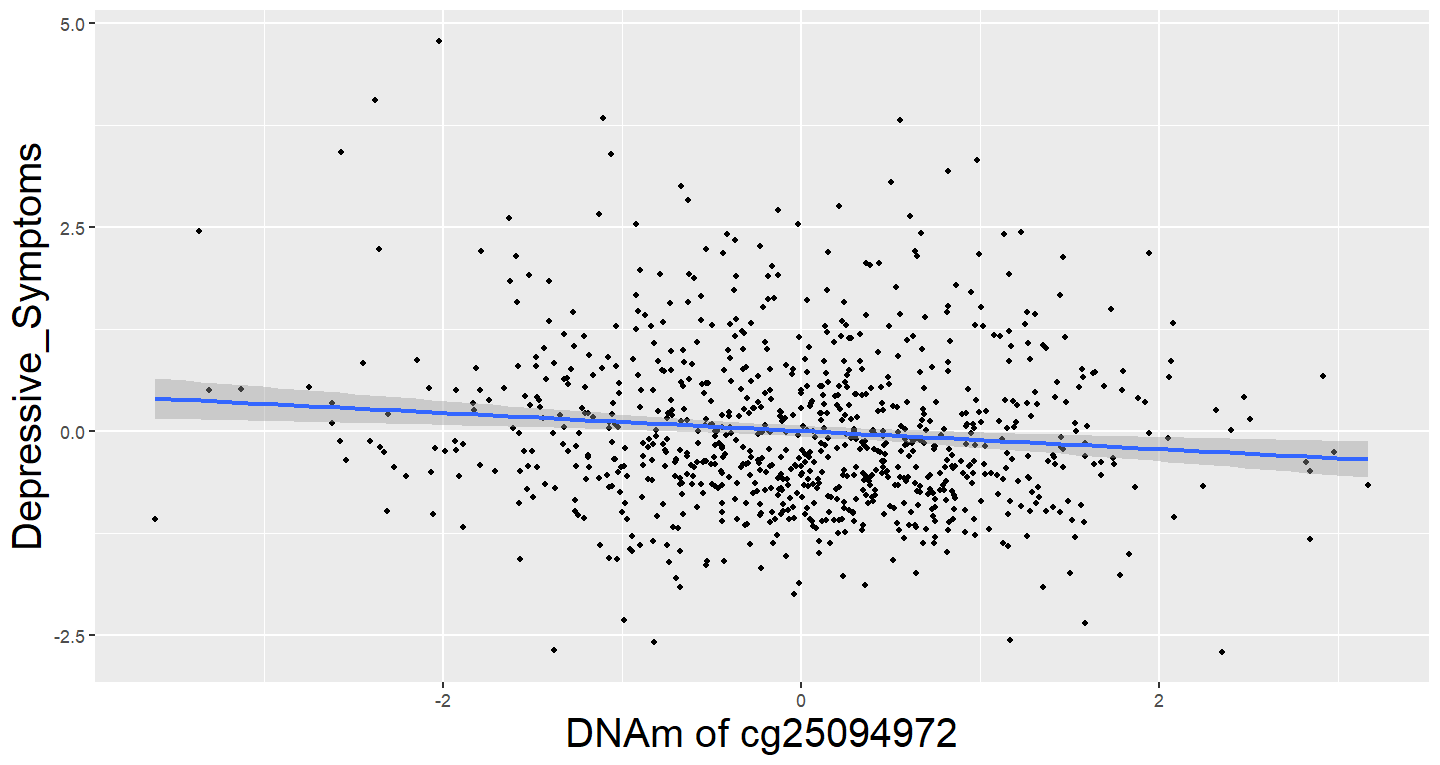

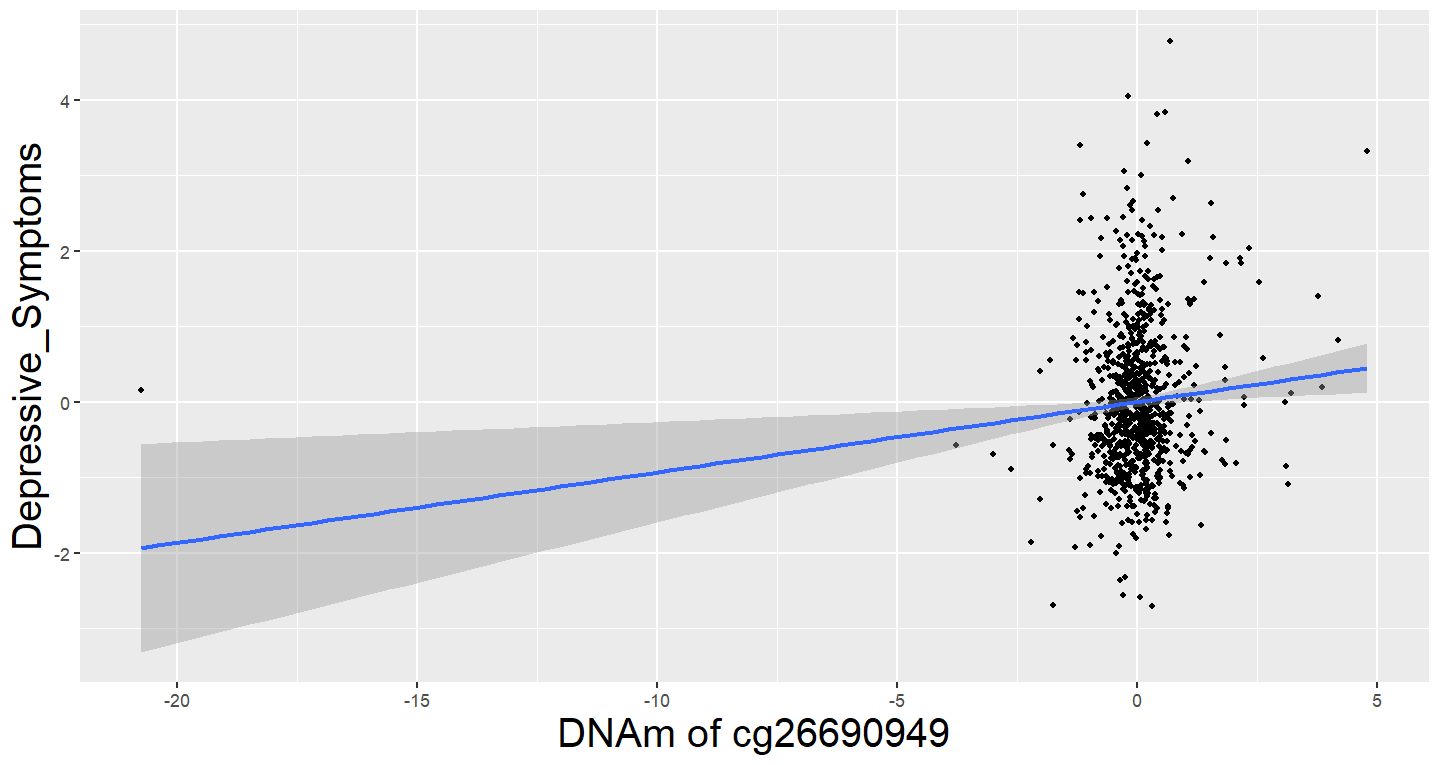

*Note.* These figures display side-by-side the associations between poor housing quality and DNA methylation of CpG sites (both measured at study enrolment); and DNA methylation and depressive symptoms (measured 3 years later) for CpG sites identified as having indirect effects.

**References**

1.	Buuren, S. van & Groothuis-Oudshoorn, K. mice: Multivariate Imputation by Chained Equations in R. *J. Stat. Softw.* **45**, 1–67 (2011).

2.	Eekhout, I. *et al.* Missing data in a multi-item instrument were best handled by multiple imputation at the item score level. *J. Clin. Epidemiol.* **67**, 335–342 (2014).

3.	Lussier, A. A. *et al.* DNA methylation mediates the link between adversity and depressive symptoms. *Nat. Ment. Health* (2024).

4.	Fan, J. & Lv, J. Sure independence screening for ultrahigh dimensional feature space. *J. R. Stat. Soc. Ser. B Stat. Methodol.* **70**, 849–911 (2008).

5.	Schaid, D. J. & Sinnwell, J. P. Penalized Models for Analysis of Multiple Mediators. *Genet. Epidemiol.* **44**, 408–424 (2020).
